# Dynamics of a slow-burn epidemic: the genomic epidemiology of poultry-associated *Campylobacter jejuni* ST474 in New Zealand

**DOI:** 10.64898/2026.10.09.26364997

**Authors:** N.P. French, D.A. Wilkinson, P.J. Biggs, A.C. Midwinter, J.M Collins-Emerson, A. Fayaz, J.C. Marshall, P. Muellner, L.E. Rogers, A. Parolin-Calarga, A.E. Mather

## Abstract

The emergence of a dominant poultry-associated strain of *Campylobacter jejuni* in New Zealand coincided with a multi-year epidemic of human campylobacteriosis. This was characterised by an increase in cases to 2006, when one of the highest national case-rates in the world was recorded, followed by a rapid decline attributed to interventions in the poultry industry. The strain belonged to an internationally rare multilocus sequence type, ST474 - a member of Clonal Complex (CC) 48 - that was highly prevalent in both poultry and human cases in New Zealand but was also identified in other samples. ST474 isolates from humans, animal reservoirs and environmental water samples were collected between 2005 and 2019 and whole genome sequenced (N=443). Isolates from a 2018/9 case-control study of campylobacteriosis in New Zealand were included for genomic comparisons with other sequence types and CCs. This presented an opportunity to examine the genomic epidemiology of a single, relatively recently emerged lineage of *C. jejuni,* and improve our understanding of drivers of emergence and persistence in a relatively isolated ecosystem.

Six subpopulations (clades) of ST474 were identified and these varied in terms of their host association, persistence, patterns of homologous recombination, and complement of mobile genetic elements, including plasmids and chromosomally integrated elements. Clade E was associated with a single poultry supplier in the initial epidemic phase and accounted for 34.5% of human cases. Clade F emerged from a declining Clade E after the peak in human cases and was also associated with poultry. The emergence of Clade F was accompanied by the loss of integrated element CJIE5 and the acquisition of multiple blocks of recombination, likely from diverse ancestral *C. jejuni* lineages, and the loss of the methionine biosynthesis operon *met*AB. In contrast, Clades A-D were detected at low frequencies in human cases over the 14-year period of surveillance and were associated with multiple hosts. This longitudinal study sheds light on the evolutionary processes that result in strain emergence and differentiation, highlighting the role of horizontal gene transfer in the evolution and adaptation of this pathogen to new niches, and the subsequent impact on public health.

## INTRODUCTION

*Campylobacter jejuni* is a globally important foodborne pathogen [1]. In 2006 New Zealand had one of the highest reported rates of campylobacteriosis in the world, triggering calls for the urgent introduction of public health measures [2]. In response to rising rates, a sentinel surveillance site was established in a region of the North Island of New Zealand in 2005 [3] and this continued until 2019, after the completion of a case-control study [4]. The data from these studies provided crucial information on the distribution of 7-gene multilocus sequence types of *Campylobacter* spp. among human cases, animal reservoirs and environmental samples. Source attribution modelling determined the most important source of human infections to be poultry, leading to the introduction of new measures in the poultry industry aimed at improving food safety [5]. Campylobacteriosis rates rapidly declined following these interventions [6].

Globally, sporadic human cases of campylobacteriosis are usually characterised by a high diversity of sequence types (STs), reflecting diversity in the sources of infection [7–12]. A remarkable feature of the New Zealand campylobacteriosis epidemic was the predominance of a globally rare sequence type, ST474, in both the human cases [5] and initially a single poultry supplier [13]. This raised questions about how such a ‘clone’ emerged in the poultry supply and became a dominant cause of human infections. The collection of isolates acquired from multiple hosts over a 14-year period, which included the poultry-associated epidemic, and the intervention in the poultry industry [6], provided a rare opportunity to study the emergence, decline and evolutionary dynamics of an important human pathogen.

Studies of the bacterial pangenome have highlighted the enormous plasticity of gene content that can occur between even closely-related bacterial strains, with some hypotheses stating that the global gene pool of bacterial genomes could be essentially infinite [14]. These characteristics of bacterial evolution have important implications for our understanding of microbial evolution and the management of emerging foodborne pathogens [15]. Bacteria can adapt to new environments rapidly, either by mutation or by the loss and gain of gene function through horizontal gene transfer (HGT). Variants of *C. jejuni* have been suggested to have adapted to the intensification and industrialisation of practices in livestock farming, particularly cattle farming, through accessory gene gain [16]. The importance of accessory genome elements for niche adaptation has also been shown to occur in *Campylobacter*, including a cattle-associated vitamin B5 biosynthesis locus within a recombinogenic genome region [17], and uptake of fucose and glucose metabolism gene clusters allowing utilisation of new substrates [18]. The result is that *Campylobacter* are omni-present in multiple livestock species, frequently contaminate the food chain and commonly cause human disease following infection [19].

We generated whole genome sequence (WGS) data from 388 isolates of *C. jejuni* ST474 from human cases, non-human animals, food products and the environment from the Manawatu sentinel surveillance site in the North Island of New Zealand between 2005 and 2018. We added 55 genomes of ST474 from other New Zealand studies, including a year-long case-control and source attribution study conducted in 2018/19 (the Source Assigned Case Control New Zealand (SACNZ) study) [4]. Other *C. jejuni* and *C. coli* genomes from CC48 and other CCs identified in the SACNZ study were included in the analysis to understand more fully the evolutionary events leading to the emergence of ST474. We use the data from this 14-year longitudinal study to explore the evolution and genomic characteristics of ST474 over this period. Previous analyses of *C. jejuni* ST474 have indicated both homologous recombination and mutation play important roles in its evolution [20]. Our extended study sheds further light on the evolutionary processes that result in spillover and niche adaptation in *Campylobacter*, particularly highlighting the significant role of mobile genetic elements, and offers insights into how future epidemic strains may arise.

## METHODS

### Origin of isolates and sequences used in this study

A total of 443 genome-sequenced ST474 isolates from New Zealand were used in this analysis. Of these isolates and sequences, 390 ST474 were generated for this study, and the remainder were generated by other studies and downloaded from public data repositories (Table 1). Sample collection dates spanned from April 2005 to March 2019. Draft genome assembly quality statistics for the 390 ST474 isolates sequenced for this study are provided in Supplementary table 1. The 390 isolates were from 378 samples; two to three colonies with different morphologies were selected and sequenced from eleven samples.

**Table 1.** Origin of genome sequenced *Campylobacter jejuni* ST474 from New Zealand analysed in this study.

| Study | ST474 |
| --- | --- |
| Unique to this study | 390 |
| Biggs et al. 2011 [20] | 1 |
| Lake et al. 2021 [4] | 43 |
| Weis et al. 2017 [21] | 4 |
| Gilpin et al 2020 [22] | 5 |
| Total | 443 |

### DNA extraction and genome sequencing

All *Campylobacter* isolates from the sentinel surveillance site that had been identified as ST474 using 7-gene MLST over the study period were revived from frozen storage and passaged twice from single colonies to ensure clonality. Isolates were grown on horse blood agar (HBA) plates, overnight in a microaerobic atmosphere at 42 ⁰C. Genomic DNA (gDNA) was extracted from an approximately 1 µl loopful of an overnight HBA culture using the QIAamp DNA Mini kit, following the manufacturer’s instructions with the RNase digestion step being performed at 37 °C for 40 mins which improved gDNA purity. gDNA yield was assessed using the Qubit HS dsDNA Assay kit, and DNA purity was assessed using both A_260_/A_280_ ratios measured by Nanodrop spectrophotometry and quantification of residual RNA using the Qubit RNA HS Assay Kit.

gDNA was sequenced on the Illumina HiSeq 2500 platform, in rapid mode generating 2 x 250 bp paired-end reads. Genomic library preps were prepared using the Nextera XT kit and adapters (Illumina), following the manufacturer’s instructions. All ST474 raw reads and assemblies generated for this analysis are available from NCBI (Bioproject ID: PRJNA1538059).

In addition to the ST474 isolates, all other isolates sequenced for the SACNZ study were included in the analysis to compare accessory gene content and identify the putative origin of loci in recombinant regions. This included sequenced isolates belonging to the same clonal complex as ST474 (CC48). A total of 1208 draft genome assemblies from the SACNZ study were downloaded from NCBI (Bioproject ID: PRJNA675916) [4]. These assemblies comprised 1052 *C. jejuni* and 156 *C. coli* whole genome sequences obtained from New Zealand isolates from humans, sheep, cattle, and poultry.

### Genome assembly and annotation

Raw Illumina read data were trimmed using a combination of Trimmomatic version 0.4 [23] to remove adapters and common contaminating sequences. Genomes were assembled using SKESA version 2.4.0 [24], and assembly statistics were extracted using seqkit version 2.1.0 [25]. Single nucleotide polymorphisms were predicted by aligning read data to the internal reference genome H1216 using Snippy version 4.6.0 [26]. This reference sequence was chosen as it had the largest genome size with one of the most contiguous assemblies within the dataset. Predicted SNPs were processed to generate a full genome alignment (FGA) using snp-sites version 2.5.1 [27], extracting monomorphic sites (option -b) and uniquely ACGT characters (option -c). All genomes were annotated using Bakta version 1.12.0 [28] and additional functional annotation of the sequences was performed using eggNOG-mapper [29] and database (version 7) [30].

### Genomic analyses: population structure, accessory genome and recombination

The population structure of the data was predicted from the FGA using the R-based implementation of the Bayesian Analysis of Population Structure software [31], rhierBAPS version 1.1.4 [32]; the software was allowed to run until it reached a local optimum with the maximum predicted number of groups set to 20. The FGA was used as the input data for Gubbins [33] under the default settings to predict mutation hotspots likely to result from recombination events, results were visualised in phandango [34] and plotted using a modified version of plot_gubbins.R ^1^. The ancestral origins of recombinant regions were evaluated by comparing allele distributions among the ST474 clades, with other clonal complexes identified in the SACNZ study. Alleles were determined using the wgMLST and cgMLST schemes following chewBBACA version 3.1.0 [35] annotation and allele assignment, and custom scripts. Maximum likelihood phylogenies of aligned allele sequences were created using the phangorn package version 2.12.1 in R [36] using the optim.pml command (model =”GTR”) and associated allele distributions by lineage were added as a heatmap using the gheatmap and ggtree commands, accessed using treedataverse version 0.0.1 [37].

Integrated elements were identified by scanning accessory genes appearing in syntenic clusters using the R package ‘dbscan’ [38], using the optimised epsilon parameter exploited by the PHASTER algorithm for prophage identification [39] and a number of genes greater or equal to five. The insertion sites for phage associated integrated elements (IEs) were determined using rtracklayer version 1.70.1 [40] to scan general feature format (gff3) files from isolates representing different clades and combinations of IEs generated using Bakta version 1.12.0 [28] and plotted using gggenomes version 1.1.3 [41]. Putative plasmids were identified using RFPlasmid version 1.2 [42] and typed using the Campy-4G scheme (https://github.com/KLIF-UU/Campy-4G). Gene content for plasmids were displayed using Geneious version R10.2.6 (https://www.geneious.com) and gggenomes version 1.1.3 [41].

### Source Attribution

Source attribution of human isolates to putative sources was performed using wgMLST allelic profiles with the asymmetric island source attribution model [43] via the islandR package (https://github.com/jmarshallnz/islandR), where mutation and recombination probabilities for each putative source were assumed to be the same. Posterior mean and variance of attribution probabilities *p̄_ij_* and *σ*(*p_ij_*) were computed for each isolate *i* to source *j* across 10,000 iterations.

### Phylodynamic modelling

A recombination masked SNP alignment was used in downstream phylodynamic analyses in order to estimate the substitution-rate, time of most recent common ancestor (tMRCA) and effective population size (N_e_). Support for a temporal signature was assessed by estimating the correlation between the optimal phylogenetic tree branch length and the sampling dates and comparing models using the sample dates with null models imposing a single sample date. These assessments were carried out using the ‘roottotip’ and ‘modelcompare’ commands in BactDating version 1.1.4 [44]. Models were run in triplicate ensuring convergence (ESS>300) for models using the time signature.

The SNP alignments were imported into BEAUti 2.5 to create an Extensive Markup Language (.xml) file for BEAST version 2.7.8 [45, 46]. The H1216 reference genome (1,683,835 bp) was used to estimate the number of invariant sites which were added as constant site weights to the .xml file. iqtree2 was used to identify the best-fitting substitution models based on the Bayesian Information Criteria (BIC) [47]. Combinations of molecular clock models including strict and relaxed clocks [48] and tree priors were trialled using nested sampling [49]. The final .xml files were run in BEAST for 50 million steps, three times with different starting seeds, before LogCombiner was used to combine the runs with a 10% burn-in. A log normal prior was placed on strict clock substitution rates with a mean in real space of 10^-5^ and a SD of 1.25 [50]. Tracer v1.6 [51] was used to visualise the results.

A maximum clade credibility (MCC) tree was generated using TreeAnnotator from the posterior distribution of trees output from a BEAST2 model using a GTR substitution model, a strict clock and a Bayesian Integrated Coalescent Epoch PlotS (BICEPS) tree prior [52]. Lineages through time were assessed using the ltt.plot function in the R package ape version 5.8-1 [53] and lineages displaying evidence of clonal expansions were identified using CaveDive version 0.1.1 [54].

## RESULTS

### Dynamics of a slow-burn epidemic

*Campylobacter* surveillance in New Zealand’s central North Island sentinel site was carried out over a 14-year period between 2005 and 2019. This included isolates collected for the SACNZ study [4]. The number of notified cases across the whole of New Zealand and in the Manawatū sentinel site are presented in Figures 1A and 1B. In the first period of the prolonged epidemic (2005-2007), prior to the major impact of the interventions in the poultry industry, ST474 was identified in up to 36% of human infections that were cultured and sequence typed (Figure 1C). After a second peak in 2009 (29% of cases) the proportion of cases that were identified as ST474 declined from 2010 to between 3 and 10% until the end of sentinel surveillance in 2019. We therefore refer to the dynamics of these ST474 infections as a three-phase slow-burn epidemic with an “epidemic phase” (2005-2007), “re-emergence phase” (2008-2011) and a “coalescent phase” (2011-2019).

**Figure 1:**
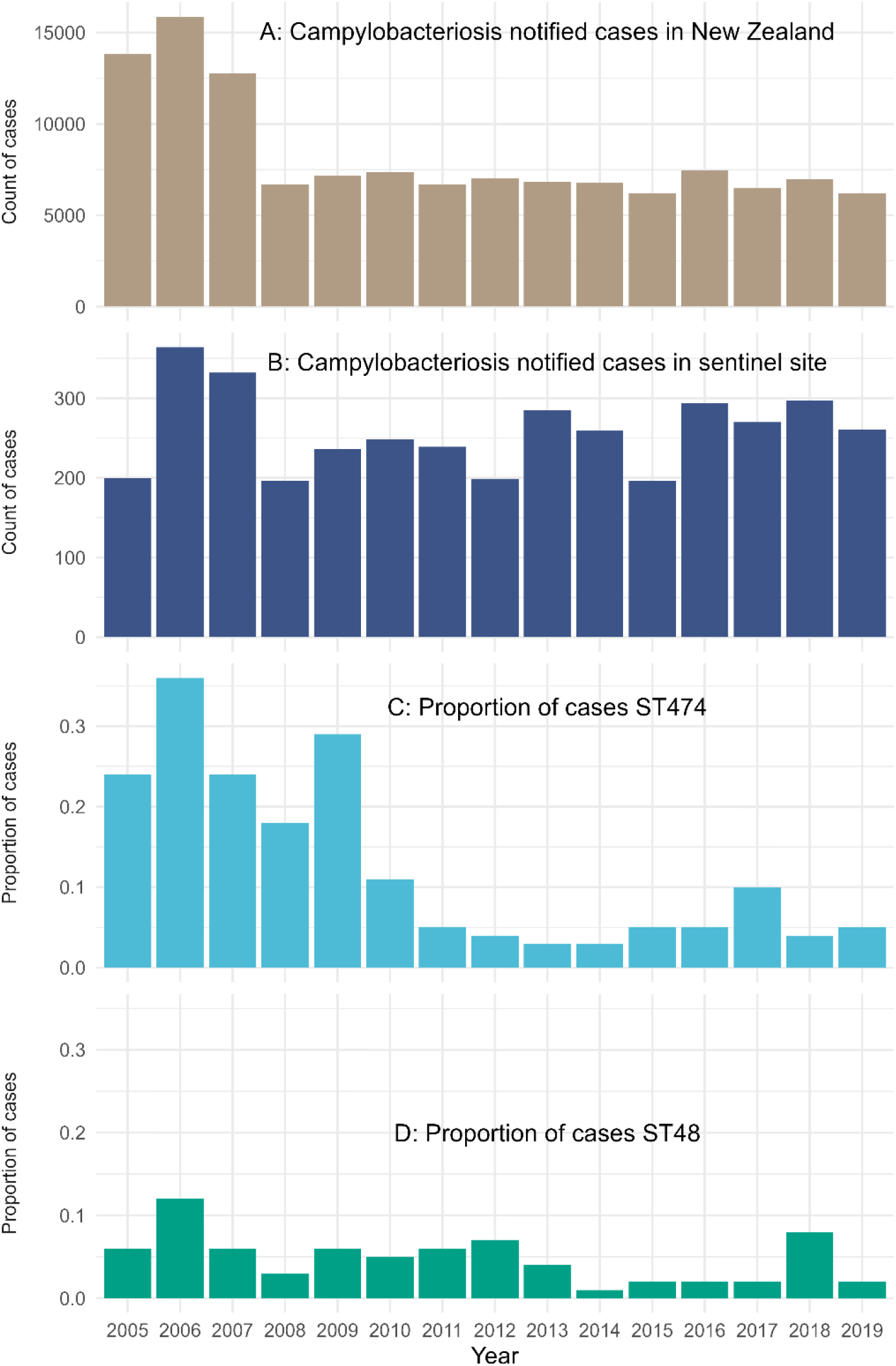
*Campylobacter jejuni* human cases between 2005 and 2019 nationally and in the campylobacteriosis sentinel site. A: Total notified campylobacteriosis cases in New Zealand. B: Total cases notified in the sentinel site. C: Proportion of cases in the sentinel site that were ST474. D: Proportion of cases that were ST48, a common sequence type from the same clonal complex as ST474 (CC48) and a single locus variant of ST474.

For comparison the proportion of human cases attributable to the related ST48 (a single locus variant of ST474) is shown in Figure 1D. The proportion due to ST48 varied from 1-12% over the time period and did not display such a marked decline after the intervention in the poultry industry.

### Genomic analysis of *C. jejuni* ST474: Population structure and host associations

Hierarchical Bayesian Analysis of Population Structure predicted a total of six main subpopulations within the ST474 isolates, labelled Clades A-F (Figures 2a and 2b). Poultry-associated Clades E and F accounted for 34.5% and 12.9% of human cases. Clade F predominated after the interventions in the poultry industry, (i.e. the re-emergence phase) between 2007 and 2012. Clade E was the dominant clade before the interventions, with 94% of human cases belonging to Clade E isolated between 2005 and 2007 (144/153). No isolates were obtained from Clade E after March 2010, despite continued surveillance efforts. No isolates were obtained from Clade F after June 2016.

**Figure 2a:**
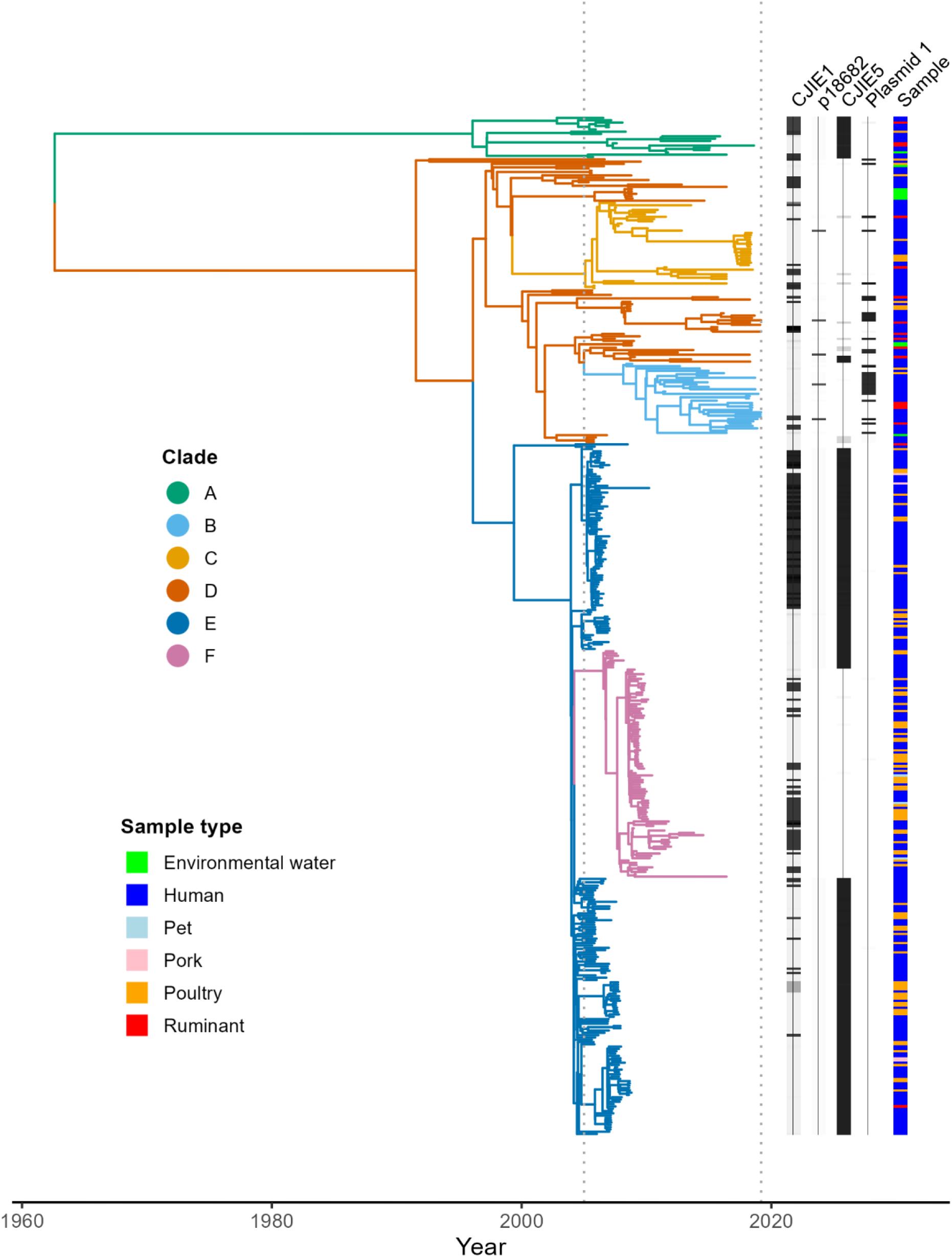
Genomic analyses of 443 *Campylobacter jejuni* ST474 isolates from New Zealand. Phylogenetic tree is a BICEPS time tree showing the estimated evolutionary histories of rHierBAPs-defined genetic Clades (A-F). The heatmaps show accessory genetic element presence/absence predictions for corresponding isolates in the phylogenetic tree and the sample type of each isolate. Lines indicate presence of each genetic element and darker shading indicates a higher percentage match (BLAST) to the chosen reference sequence for each genetic element.

**Figure 2b.**
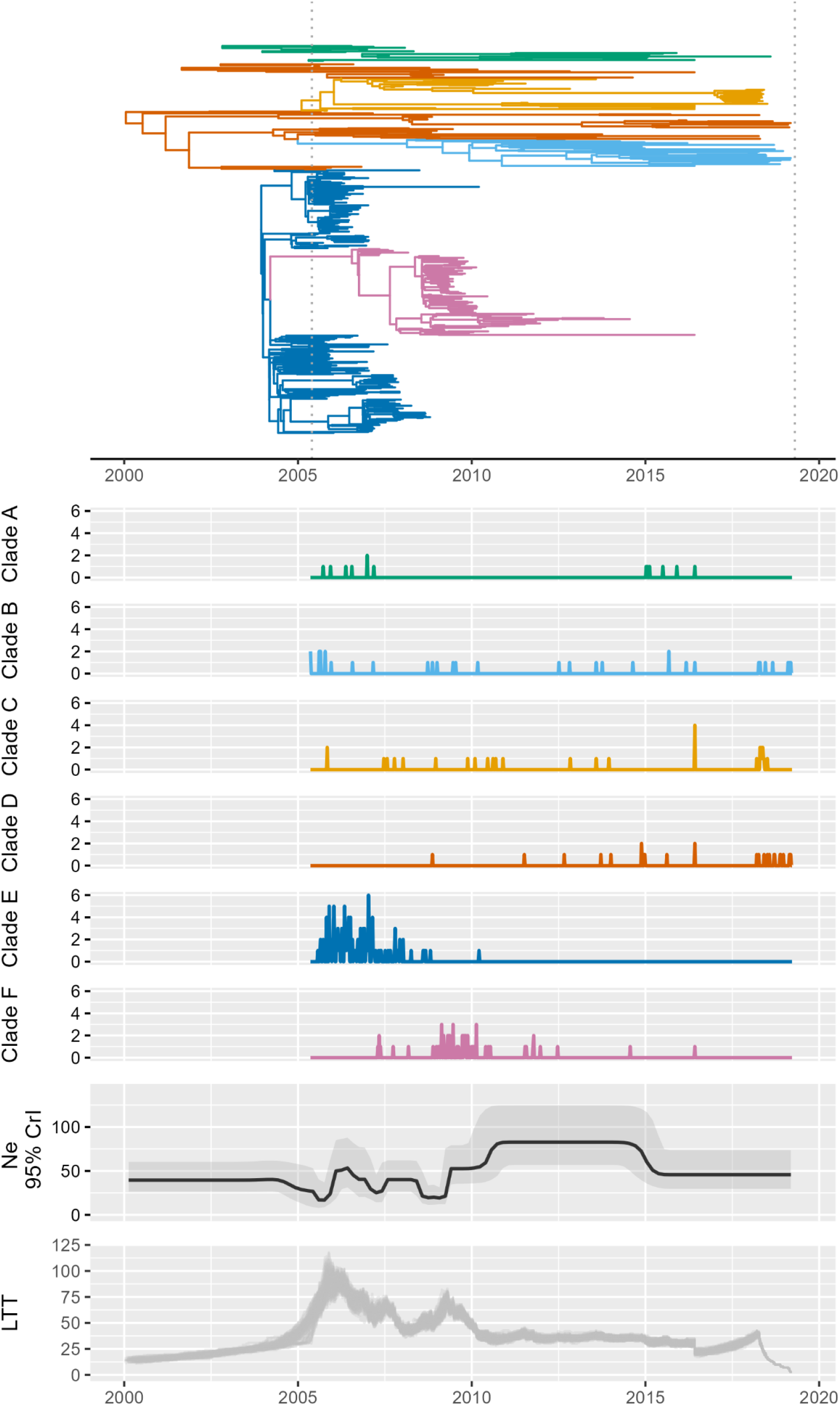
Time series of notified human cases from each Clade (A-F) and the estimated effective population size (N_e_) (black) with 95% HPD intervals (shaded) and lineages through time (LTT) plot including data from 100 trees sampled from the posterior distribution of trees. The extracted time tree for the period 2000 to 2019 is shown above the time series plots.

Non-human isolates belonging to Clade E were predominantly obtained from point-of-sale chicken samples (n=43, 87.8%) but were also identified in pork and beef meat products and in one sample of environmental water. Amongst Clade F non-human isolates, 92.9% (n=39) originated from point-of-sale chicken samples, with remaining isolates coming from samples associated with companion animals [55]. Asymmetric island model attribution results indicated that poultry were the most likely source of infection for all cases associated with both Clades E and F, with strong associations with poultry isolates collected before and after the intervention accounting for the majority of human cases isolated over those same time periods (Figures 3a and 3b).

**Figure 3.**
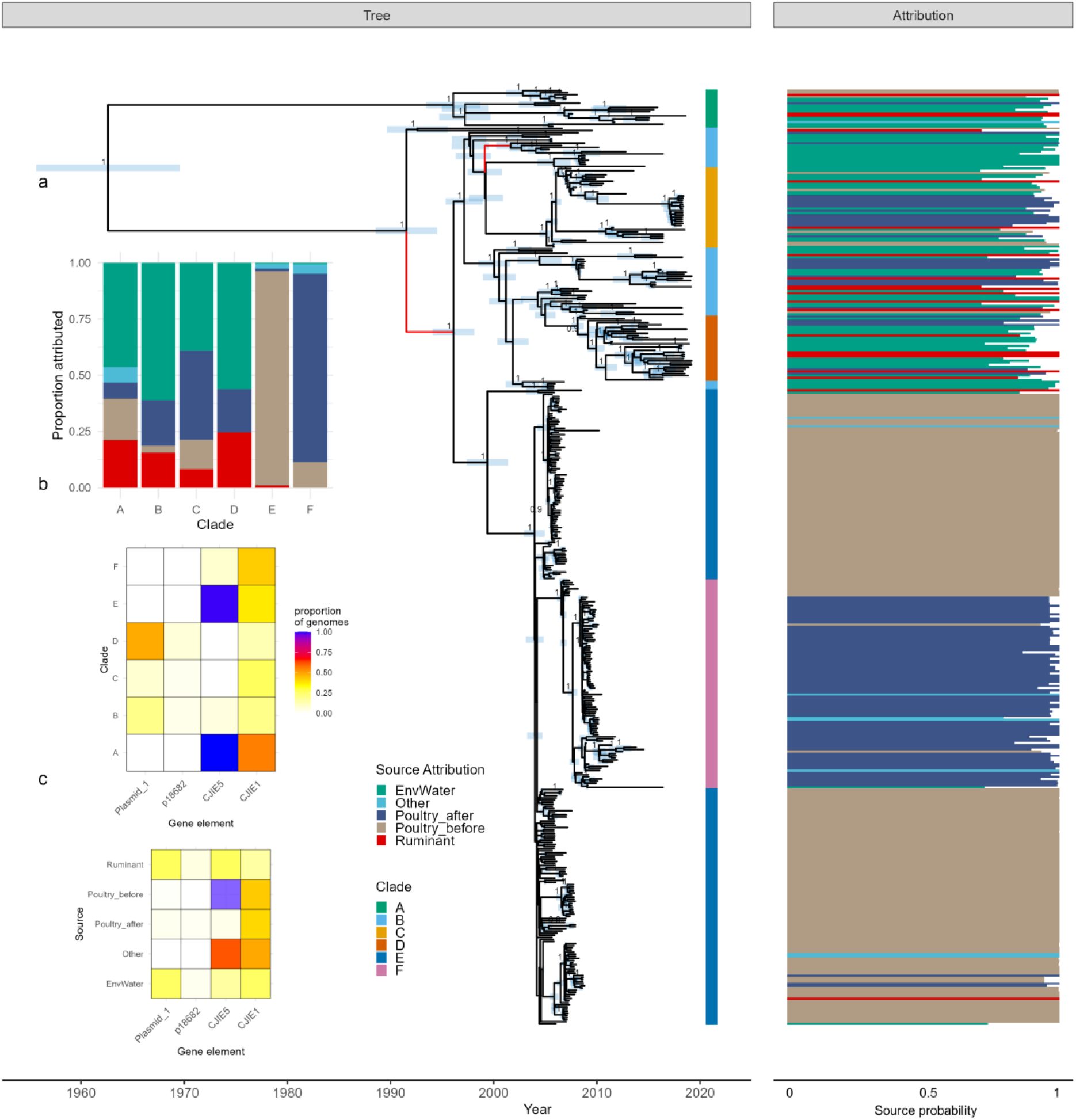
The tree (a) represents the BEAST time tree showing the estimated 95% HPD intervals for the date of each node (light blue bars) and the estimated posterior probabilities for each node >0.9. The two red internal branches are strongly identified as periods of clonal expansion using CaveDive. The clades are indicated in the bar to the right of the tree. The attribution heat map shows the source attribution results for each isolate. Non-human isolates are assigned to their source with probability 1.0. The source of human isolates is estimated from the asymmetric island model. Colour indicates the most likely source to which each case was attributed, and bar width indicates the probability of attribution to that source (probability 0-1). Inset b summarises the proportion of human isolates attributed to each source for each clade. Inset c indicates the proportional presence of different accessory genetic elements in genetic clades (top) and isolates attributed to different sources (bottom), using the same scale.

All remaining genetic subpopulations of ST474 (Clades A-D) were detected at lower frequencies in human cases throughout the entire 14-year period of surveillance in both non-human samples and human cases (Figure 2b). Further, the majority of isolates collected during the coalescent phase, including all isolates collected after 2016, belonged to Clades A-D. Source attribution results indicated a relatively higher contribution from non-poultry sources (ruminants and environmental water) to human cases belonging to Clades A-D (Figure 3b), compared to the predominantly poultry-associated Clades E and F.

Clade A diverged near the root of the time tree and was associated with long branches connected to the other clades. As this clade was the most divergent and was persistently identified in low numbers throughout the study, it was referred to as the ‘basal clade’.

### Mobile genetic elements: integrated elements CJIE5 and CJIE1

The acquisition or loss of two integrated genetic elements was associated with the evolutionary history of ST474, and the emergence of the epidemic and re-emergence of clades E and F. The first was a variant of integrated element CJIE5 first identified in ST677 isolates from human cases in Finland [56]. In ST474 isolates this was an approximately 24kb region (Figures 2a, 3c, 4 and Supplementary Figure S1). The sequence showed 100% query coverage and 99.7% identity to a CJIE5 extracted from a ST677 genome (Genbank: CP010461) [56]. Bakta identified two endonucleases in the ST474 CJIE5 sequences: a DNA/RNA non-specific endonuclease and a restriction endonuclease adjacent to a KAP NTPas domain-containing protein and a DNA methyltransferase (Figures 4 and Supplementary Figure S1). Although CJIE5 contained a 314bp phage protein gene and a site-specific integrase, it lacked other phage-associated genes.

**Figure 4:**
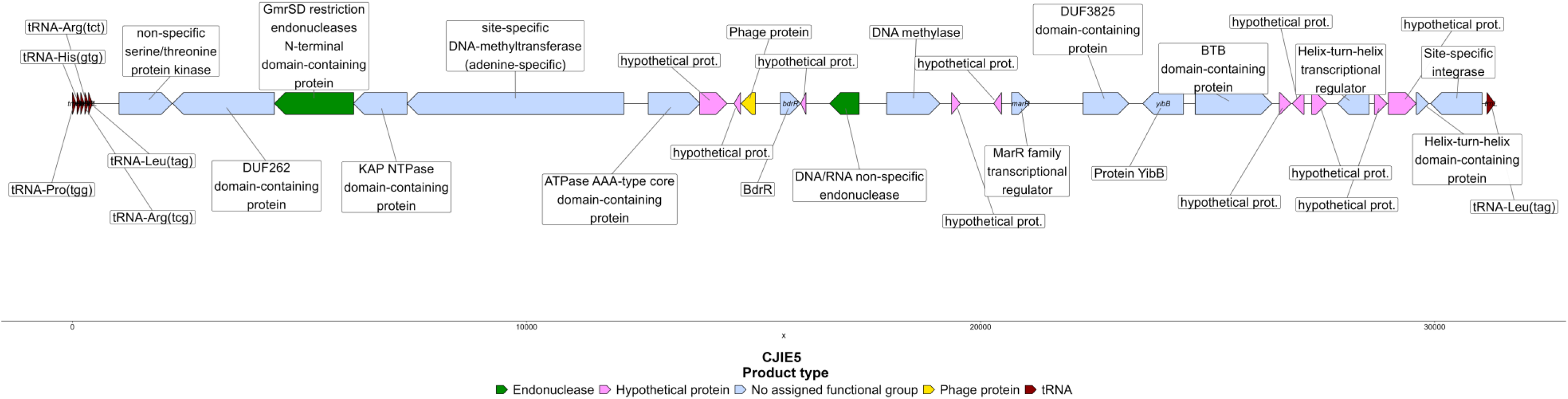
Schematic representation of integrated element CJIE5 identified in ST474 *Campylobacter jejuni* isolates from this study annotated using Bakta version 1.12.0 [28].

CJIE5 was detected in all ST474 isolates in Clade A (the clade that branched close to the root node of ST474), and the majority of isolates in Clade E (98.5%, 200/203). In contrast, it was rarely detected in clades B, C and D and was absent in all Clade F isolates sampled after 4/3/2008 (a sub-clade named Clade F_REC, see the section below on recombination). It was also absent in all isolates identified as ST48.

The second major integrated element, a variant of CJIE1, is an approximately 40kb prophage sequence (Figures 2a, 3c and Supplementary Figure S1) that was identified in 34% (151/443) of ST474 isolates in all clades. It was present in all isolates belonging to ST48 in the SACNZ dataset and was more similar to CJIE1 than the other integrated elements identified in the reference genome RM1221 (CP000025.1) [57], with 98% identity but only 61% query coverage. According to a BLAST analysis, it was most similar to a prophage homologue identified as a putative CJIE1 in *C. jejuni* subsp*. jejuni* 00-2425 isolated from a waterborne outbreak of campylobacteriosis in Canada (CP006729.2) [58, 59] with 95% coverage and 99.97% identity. In addition to multiple phage-associated genes, CJIE1 contained a gene encoding a bacteriocin and a deoxyribonuclease (DNase) with 100% query coverage 99.9% identity to the DNase identified in reference genome RM1221 (CP000025.1).

### Integration sites for CJIE5 and CJIE1

CJIE5 was located in a variable region bounded by a glycoprotein gene, *cgp*A, and a conjugal transfer protein, *traF*, (named *cgp*A-*traF* in Supplementary Figure S2a, Supplementary Table S2). In isolates without an IE in this integration site, the region contained an ATP-dependent DNA ligase, five tRNAs (Pro(tgg), His(gtg), Arg(tcg), Arg(tct) and Leu(tag)), and two genes encoding hypothetical proteins (117bp and 438bp). CJIE5 was consistently inserted between the 85bp tRNA-Leu(tag) and the 116bp hypothetical protein gene in the *cgp*A-*traF* insertion site. In all CJIE5 identified in ST474 there was 177bp gene annotated by Bakta as a tRNA-Leu(tag) adjacent to the right-end junction (the 116bp hypothetical protein gene). Alignment of the smaller 85bp tRNA-Leu(tag) with the putative 177bp gene showed 100% sequence similarity for the final 53 nucleotides of the two genes, indicating a potential region of disruption of a functional tRNA-Leu(tag) with multiple poly A and poly T tracts (Supplementary Figure S3).

When present in isolates from Clade E, the full-length integrated element CJIE1 was also located in the integration site *cgp*A-*traF* and was inserted within CJIE5, between a DUF262 domain-containing protein gene and the GmrSD restriction endonuclease gene (Supplementary Figure S2b). An approximately 30kb truncated variant of CJIE1 was present in 5 isolates in a sub-clade of Clade E (visible as a darker grey block in the lower half of Clade E in Figure 2a). This was also located in the *cgp*A-*traF* integration site inside CJIE5, upstream of the 177bp tRNA-Leu ‘pseudogene’, in two of the 5 isolates (H609 and T124 in Supplementary Figure S2c). The integration site *cgp*A-*traF* gene contents are summarised in Supplementary Table S2. The location of CJIE1 in clades A-D and F varied both within and between clades and was identified in at least three locations in addition to the *cgp*A-*traF* integration site.

### Mobile genetic elements: plasmids

We identified two putative plasmids associated with Clades B, C and D. The first was an approximately 28kb putative plasmid, designated “plasmid1”, which was identified in 6.5% (29/443) of ST474 isolates, and only in Clades B, C and D (Figure 2a, 3c, 5 and Supplementary Figure S4). This likely-conjugative plasmid contained genes for both the conjugative apparatus and plasmid replication, as well as genes for enzymes linked to amino-acid and ketone metabolism (Figure 5 and Supplementary Figure S4). It was typed as an M5_R2_S1_V3 according to the four-locus Campy-4G scheme (https://github.com/KLIF-UU/Campy-4G) and shows genetic similarity to plasmids identified in other *C. jejuni* and *C. coli* isolates from New Zealand, the USA and Europe (from at least ten different STs). A detailed analysis of plasmids in the SACNZ study revealed 157 isolates from 30 different STs had plasmid markers M5_R2_S1_V3 in the same contig (Aline Parolin Calarga, pers comm).

**Figure 5:**
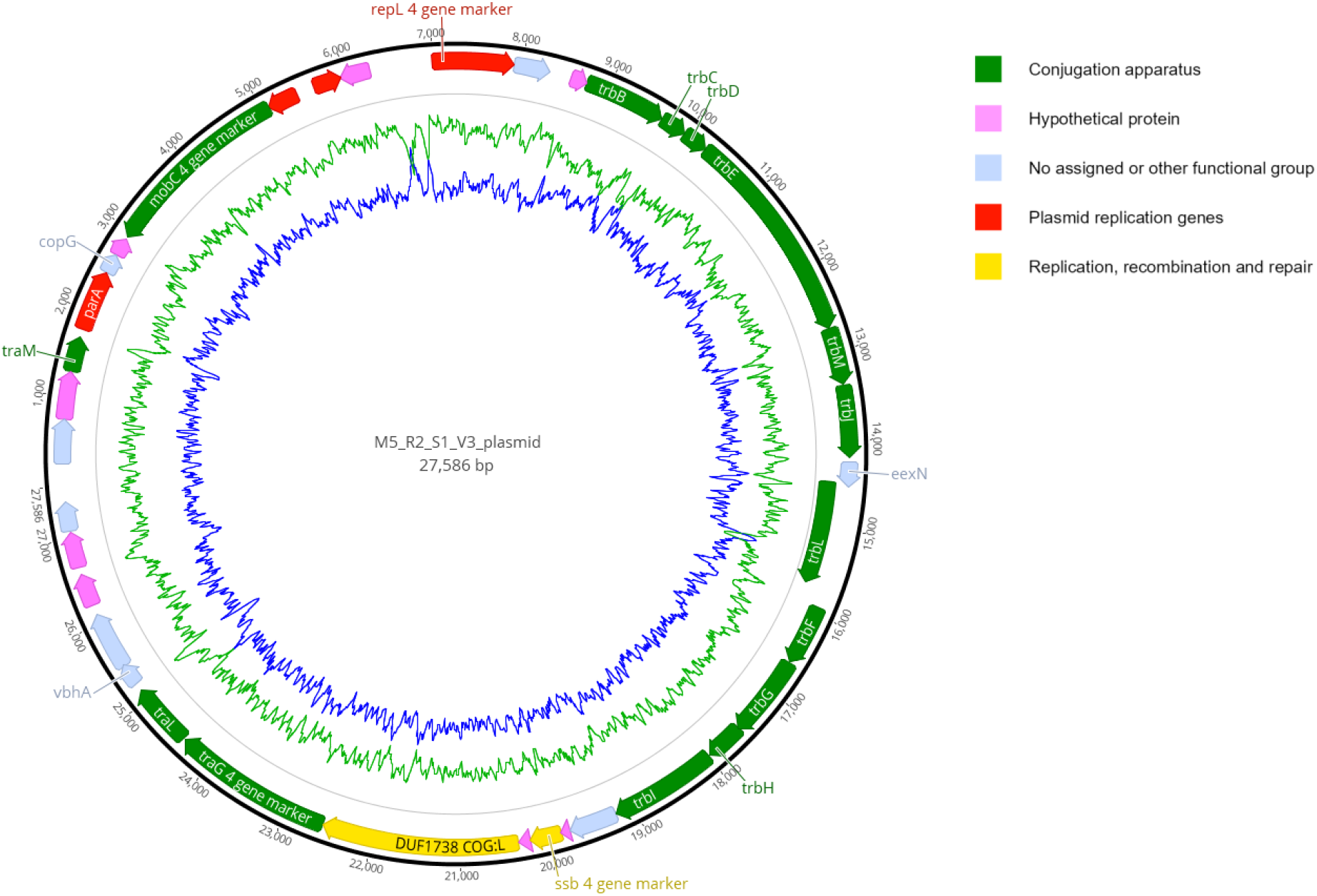
Schematic representation of plasmid1 the most prevalent plasmid in ST474 *Campylobacter jejuni* isolates from this study. The markers used in the Campy-4G typing scheme are indicated and this plasmid was typed as M5_R2_S1_V3. The plasmid was annotated using Bakta [28] with additional information on COGs provided by eggNOG [29]. GC (green) and AT (blue) graphs are shown in the inner rings. Further details of this plasmid are provided in Supplementary Figure S4.

The second was an approximately 47kb plasmid typed as an M4_R59_S3_V1 plasmid with nearly 100% genetic identity to conjugative plasmid p18682 described in a *C. jejuni* isolated in the Vellore region of India in 2019 (GenBank CP050684.1). BLAST results suggested that similar variants of this plasmid occur frequently in *C. jejuni* and *C. coli* isolates (at least 13 different STs, all carrying the *tet*O gene) from the USA, Germany, Netherlands, and different countries in Asia (% identity > 96% and cov > 77%). The *tetO* gene encodes an efflux pump that confers resistance to tetracycline-based antibiotics. This plasmid was identified in 0.9% (4/443) of ST474 isolates, all belonging to Clades B, C and D (Figures 2a and 3c) and 52% (43/83) of the ST48 isolates from the SACNZ study. A detailed analysis of plasmids in the SACNZ study revealed 49 isolates from 21 different STs had the plasmid markers M4_R59_S3_V1 in the same contig (Aline Parolin Calarga, pers comm).

A third plasmid was identified in a single human isolate in Clade B. This was a 36kbp plasmid typed as M10_R10_S2_V2 (Supplementary Figure S4). The presence of these three plasmids in the (non-poultry associated) Clades B, C and D suggest they may have been independently and horizontally acquired from environmental sources on multiple occasions. They were the three most prevalent plasmid types in isolates from the SACNZ study (Lake *et al*. 2021, pers comm Aline) and were identified in multiple STs and both *C. jejuni* and *C. coli* isolated from New Zealand samples.

### Homologous recombination

The proportion of the genome estimated to be contained within recombinant regions was higher in Clades B-F compared to the basal Clade A (Figure 6). Recombinant regions increased significantly (P<0.001) over time for Clades B, D, E and F, and decreased significantly (P=0.03) over time for Clade A. The trend for Clade C was not significant (P=0.13) (Supplementary Figure S5.)

**Figure 6:**
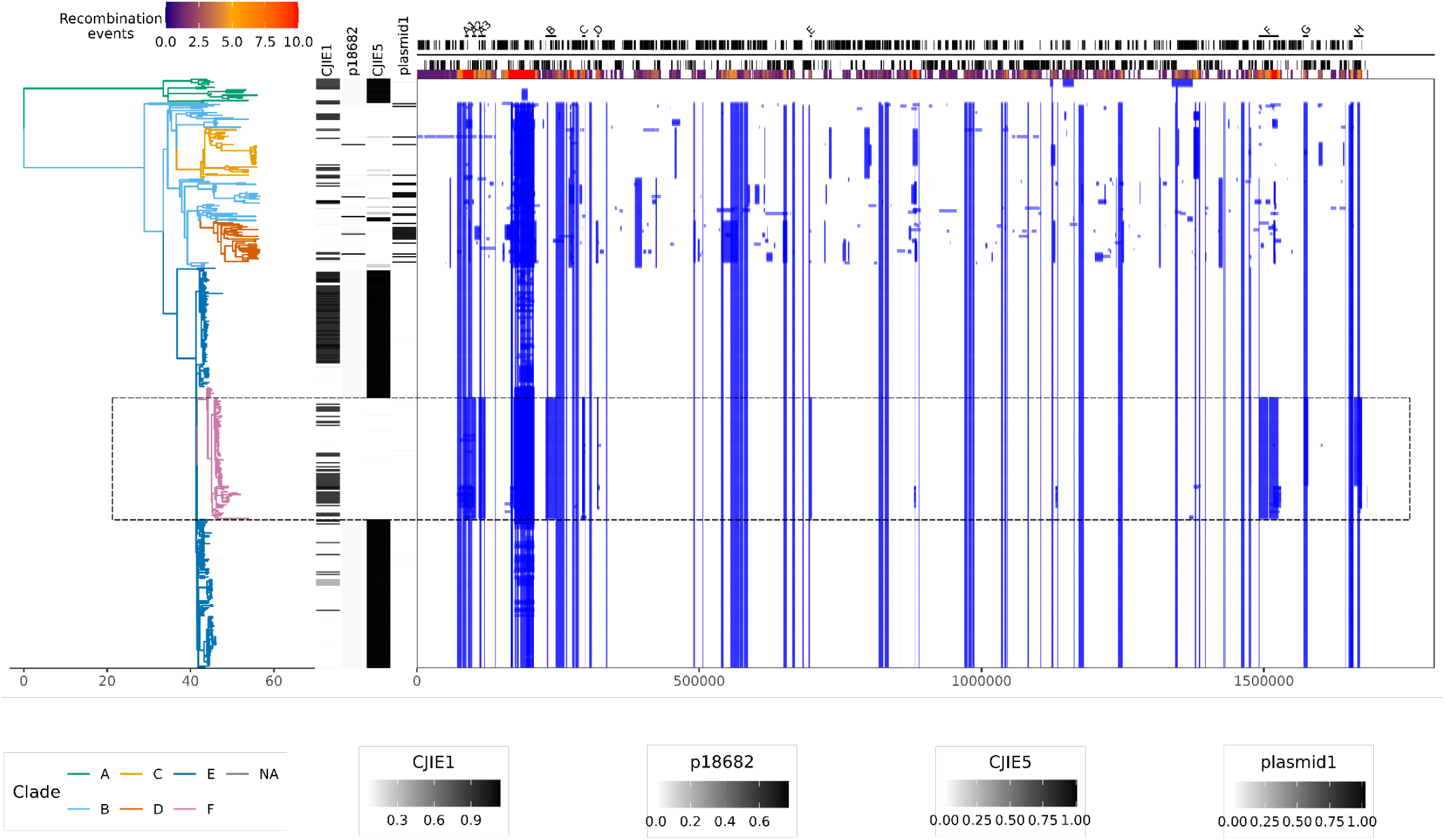
Analysis of the evolutionary history and recombination events for *Campylobacter jejuni* ST474 isolated from New Zealand. The panel on the left shows the BEAST2 time tree with branches coloured by Rhierbaps clades. The time scale is in years from the time of most recent common ancestor. The heat map to the right of the tree shows inferred recombination events shared through common ancestry in blue. The annotation based on internal reference isolate H1216 is shown at the top of the figure, with the coordinates at the bottom of the figure. Regions associated with recombination in re-emergence Clade F_REC (magenta-coloured branches, dashed black box) are highlighted above the annotation.

The largest and most distinctive recombinant regions were associated with Clade F, the re-emergence clade. These are highlighted in Figure 6 (regions A-G) and described in detail in Supplementary table 3. Multiple recombinant regions were evident in a group of Clade F isolates collected after 4/3/2008 (referred to as Clade ST474_F_REC contained within the black box in Figure 6). These included genes and operons associated with capsular biosynthesis and lipooligosaccharide (LOS) production, flagellar assembly and modification (FM), heat shock protein (HSP) production and regulation and iron transport – these are genes involved with immune evasion and virulence (FM and LOS) and survival in different environments (HSP and iron transport) (Figure 6). A detailed analysis of the alleles found in these recombinant regions revealed evidence of evolutionary events prior to the emergence of Clade F_REC, including the acquisition of recombinant tracts from different ancestral lineages.

### Recombinant regions in Clade F_REC: alleles, ancestry and the *met*AB locus

The largest of the recombinant regions identified in Clade F_REC comprised discontinuous blocks A1-A3 (∼37Kb), and contained genes involved in pseudaminic acid biosynthesis and legionaminic acid pathways essential for O-linked flagellar glycosylation [60]. We examined alleles for seven contiguous genes in Region A3 involved in the pseudaminic acid pathway (*pse*I, *pse*A, *his*H1, *his*F2, *pse*H, *pse*G and *pse*F). The majority (91%, 319/352) of isolates from Clades A-E and the non-recombinant Clade F_NR isolates, had indistinguishable nucleotide alleles in all seven genes within this region. These alleles were prevalent in the wider SACNZ dataset, and common in multiple other STs and CCs, including STs belonging to CC48 (Figure 7 and Supplementary Figure S6).

**Figure 7.**
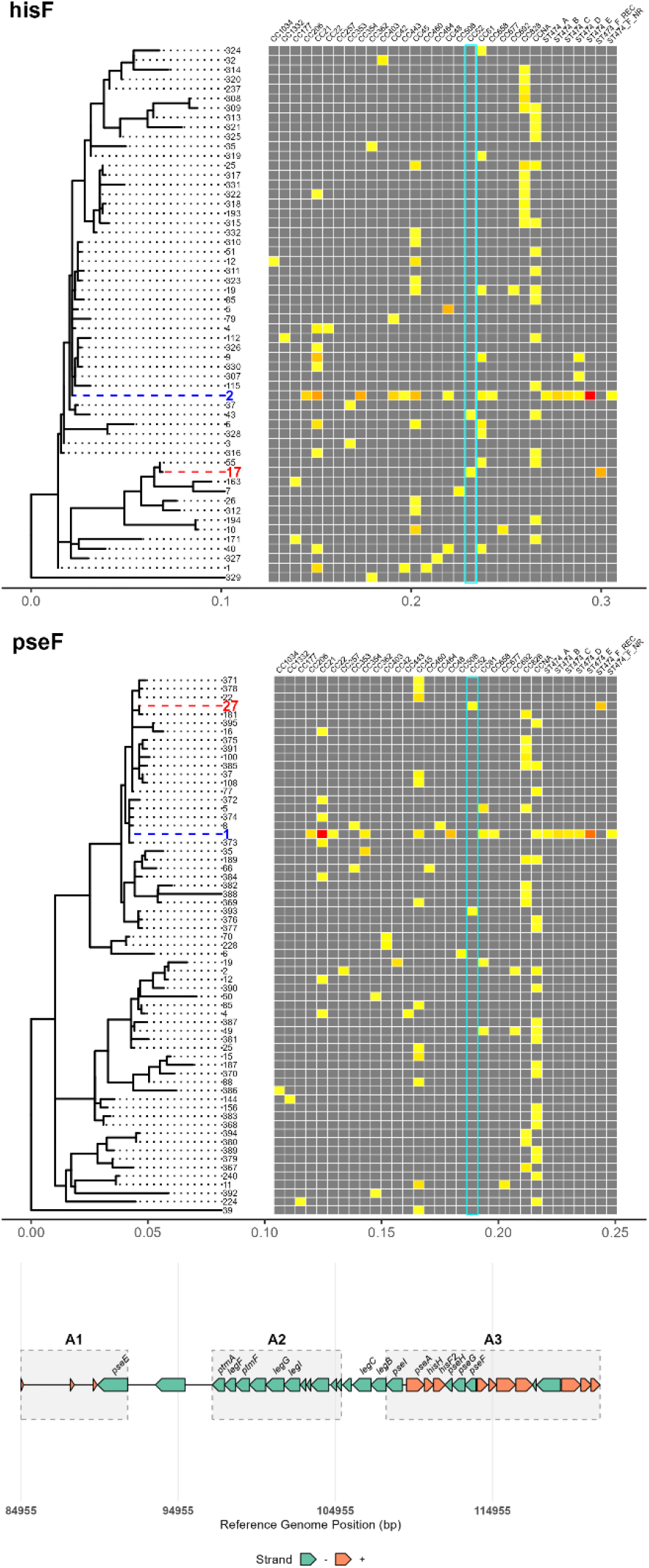
Maximum likelihood phylogenies of two genes in the pseudaminic acid biosynthesis pathways (*his*F2 and *pse*F), involved in flagellar glycosylation in *Campylobacter* spp. The blue tip label corresponds to the allele most prevalent in ST474 Clades A-E and Clade F_NR. The red tip label corresponds to the allele identified in Clade F_REC. The heatmap shows the alleles present in each ST474 clade and Clonal Complexes identified in the SACNZ study, with grey representing not present, red being the most frequent and yellow being the least frequent. The cyan box highlights the alleles identified in CC52, which shares alleles with ST474 Clade F_REC. The coordinates of genes in recombinant Regions A1-A3 are shown (reference genome H1216). The alleles in recombinant Clade F_REC were phylogenetically distinct from those in isolates in other ST474 isolates. Their similarity with alleles from different CCs indicates an importation of genes from an isolate or isolates belonging to a divergent lineage.

In contrast, the alleles for these seven genes in isolates belonging to recombinant Clade F_REC were located in a distinct branch of the phylogenetic tree, indicating different evolutionary ancestry, most likely attributable to recombination with a divergent lineage of *C. jejuni* prior to the emergence of Clade F_REC (tip labels highlighted in red in Figure 7). Isolates from the divergent CC52 were associated with the recombinant alleles identified in Clade F_REC isolates, sharing identical *his*H1, *his*F2, *pse*H, *pse*G and *pse*F alleles. In the remaining two loci, *pse*I and *pse*A, the alleles in Clade F_REC isolates were identical and unique to this clade. For illustration, the phylogeny of *his*H1 and *pse*G and the relationships between alleles, clades and CCs for these two genes are shown in Figure 7. All seven genes investigated in Region A3 are included in Supplementary Figure S6 (CC52 alleles are highlighted in cyan boxes).

The first half of recombinant region F (∼36Kb, Figure 6 and Table 2), contained genes that were more likely to share alleles with isolates belonging to CC354, including genes involved in amino acid and cofactor biosynthesis (*que*C, *ilv*D, *glt*D and *glt*B), and stress response and protein stability (*dsb*I, *rrc*). Genes in the second half of region F were more likely to share alleles with isolates belonging to CC21, including genes involved in DNA replication and repair (*dna*N, *dna*A and *ruv*C), and flagellar biosynthesis (*flg*E2).

Examination of the region between *flg*E2 and *que*F revealed the absence of methionine synthesis genes *met*A and *met*B in all 91 Clade F_REC isolates. These were present in 343/352 of remaining ST474 isolates, including Clade F_NR (Supplementary Figure S7). The most common allele for *flg*E2 identified in 76/91 Clade F_REC isolates was also found in 40/301 CC21 isolates, all belonging to ST190. In contrast to the Clade F_REC isolates, the *met*AB operon was present in all 40 ST190 isolates from the SACNZ study.

The evidence of different recombinant regions containing alleles from several different CCs is consistent with the acquisition of multiple tracts from different ancestral lineages prior to the emergence of Clade F_REC.

### Phylodynamic analysis

Root-to-tip analysis strongly supported the existence of a time signal (R^2^ value 0.55, P<0.0001, x-intercept = 1958-1959) (Supplementary Figure S8) and there was stronger support for a model with tip dates corresponding to the date of isolation than a model with uniform simulated dates (converged model with sampled dates has a DIC=3496.00 and the null model with identical dates has a DIC=3640.43).

Nested sampling provided very strong support [61] for a strict clock over a relaxed clock. We therefore present the results from a BEAST analysis using a strict clock, a GTR substitution model and a BICEPS tree model. The estimated tMRCA of all ST474 isolates was ∼1963 (95% HPD 1956-1970) (Figure 3a). Clade A, the non-poultry (predominantly ruminants and environmental water) associated ‘basal’ clade, and Clades B-F diverged from each other at the root of the time tree. The tMRCA for poultry associated Clade E was estimated to be ∼1999 (95% HPD 1997-2001) and for re-emergence Clade F was ∼2007 (95% HPD 2006-2007). These estimates and the estimated total effective population size (N_e_) were robust to the choice of substitution model.

Estimates of N_e_ revealed evidence of an increase just prior to the peak of human cases associated with poultry-associated epidemic Clade E in 2006 followed by a decline during the intervention period to 2008 (Figure 2b). The N_e_ was estimated to have increased again during the re-emergence phase and was associated with the increased detection of poultry-associated Clade F. The lineages through time plot reflects the dynamics of the poultry associated Clades E and F (Figure 2b). The N_e_ values were estimated to remain relatively high during the coalescent phase, although these estimates were accompanied by relatively wide 95% HPD intervals.

Analysis of tree structure and clonal expansion using CaveDive provided strong evidence for two clonal expansion events (Figure 3 and Supplementary Figures S9 and S10). The first coincided with the emergence of Clade B and the cluster of isolates including the majority of isolates from Clade B and all isolates in Clades C to F (Figure 3a long red branch). This branch was estimated to have expanded around 1998. The second expansion event was estimated to be prior to the emergence of a small subgroup of isolates from human cases and poultry in Clade B.

## DISCUSSION

*Campylobacter jejuni* ST474 has had a major impact on public health in New Zealand over an extended period as a result of its prominence as a foodborne pathogen. It was initially identified as a dominant strain during the large and growing epidemic of campylobacteriosis in 2005, when 7-gene MLST was used for the first time. Despite being internationally rare [62], it was the dominant strain in the North Island campylobacteriosis sentinel site, and common in other New Zealand locations [62], including a South Island region used for evaluation of the representativeness of the sentinel site [3]. Using low-resolution 7-gene MLST, source attribution modelling identified it as strongly associated with poultry [5, 13], and this finding led to interventions in the poultry industry that was followed by a marked decline in human cases [6].

The advent of whole genome sequencing provided an opportunity to study the evolution and population dynamics of this pathogen in greater detail. A study conducted in 2011 compared two New Zealand ST474 isolates using 36bp read Illumina technology [20]. Despite the use of relatively short read sequences, this study identified the importance of horizontal gene transfer in the evolution of this pathogen and concluded that both mutation and recombination are important with the latter contributing to greater divergence; a finding that has been supported by other studies of *Campylobacter* spp. [63–66].

Our analysis of 443 ST474 genomes from isolates collected over a period of 14 years, spanning the rise and decline of the epidemic in New Zealand, provided a unique opportunity to conduct a detailed investigation of the evolution and emergence of this pathogen. The evolutionary trajectory was characterised by divergence into at least six distinct populations (Clades A-F). The study provides evidence of a ‘basal clade’ (Clade A), source-attributed to non-poultry reservoirs, that emerged from the root, with an estimated date of common ancestor of ∼1960. Isolates belonging to this clade were identified throughout the period of study. The clade is characterised by two integrated elements (IEs) and relatively low homologous recombination compared to all other clades; a feature that may be attributable to the presence of at least one of the IEs [67]. This clade may therefore represent a relatively stable ancestral population that was the origin of all other clades, including the poultry-associated Clades E and F. Given the close geographical proximity of different livestock species, including the highest densities of both cattle and poultry in the Waikato region of North Island [68, 69], emergence may have been characterised initially by a crossover from environmental reservoirs, potentially from ruminants into the poultry supply, followed by widespread transmission in the poultry sector [70, 71]. In a separate outbreak, the rapid spread of an antimicrobial resistant *C. jejuni* sequence type in the New Zealand poultry supply (ST6964) further illustrated the potential for local spread with major consequences for public health [71].

Analysis of clonal expansion of individual lineages provided evidence of an exponential increase in the effective population size N_e_ in a branch containing the majority of isolates in Clade B, and all isolates in Clades C-F. This may be attributed to the colonisation of new niches following the divergence from the basal Clade A. The estimated N_e_ of the whole population shows evidence of fluctuations during the intervention period, just prior to the peak in cases dominated by poultry-associated epidemic Clade E and the emergence and apparent disappearance of poultry-associated re-emergence Clade F. These fluctuations are consistent with amplification in poultry flocks leading to the peak in human cases, a subsequent decline coinciding with the intervention in the industry, followed by a second period of amplification as Clade F_REC emerges. In contrast to the rapid emergence and decline of poultry associated Clades E and F, the other clades appear more stable and persist throughout the period of study.

HGT is a key feature of the evolution of ST474 in NZ. The emergence of poultry clades was characterised by the acquisition and loss of IEs and conjugative plasmids, indicating roles for transduction and conjugation. However, the most striking feature was arguably the role of natural transformation, with evidence of extensive recombination in all clades except the ‘basal’ clade A. In clades B-F there was evidence of an increase in the proportion of the genome in recombinant regions (Supplementary Figure S5), with the highest rate of increase, and the highest proportion of the genome that was identified as recombinant was evident in re-emergence Clade F. A detailed examination of re-emergence Clade F revealed a relatively high proportion of the genome comprised multiple recombinant regions, and this increased markedly after March 2008 when a subclade (Clade F_REC) emerged and became dominant in poultry and human cases. Human cases and poultry detections declined for Clade F_REC over the subsequent two years and after June 2016 this clade was no longer detected, signalling the possible extinction of both epidemic Clade E and re-emergence Clade F. We investigated the possible genomic reasons for the emergence and apparent extinction of these clades.

Homologous recombination occurs frequently in *Campylobacter jejuni* and *C. coli* and many studies have examined the relative rate (relative to mutation) and the absolute frequency of recombination events in these two species [63–66]. It is considered a key mechanism to acquire horizontally-transferred genomic material from other related bacteria that can facilitate survival in new niches [72]. The earlier study of recombination in ST474 revealed evidence of extensive recombination originating from multiple ancestral lineages [20]. The emergence of Clade F_REC from Clade E and Clade F_NR was associated with the acquisition of multiple recombinant tracts throughout the genome, most likely through natural transformation and the uptake of naked DNA from the host environment.

A comparison of alleles in multiple recombinant regions with a dataset including multiple STs and CCs from the New Zealand SACNZ study [4] provided evidence that these recombinant regions were acquired from at least three highly divergent *C. jejuni* lineages. This was most notable in Clade F_REC which could be described as a ‘mosaic’ clade comprising a backbone derived from Clades E and F_NR, with HGT from CC52, CC354 and CC21 lineages. The emergence of this lineage was associated with the loss of CJIE5 and the accumulation of multiple recombinant tracts, including a large tract (regions A1-A3) containing genes encoding the pseudaminic acid pathway (*pse*I*, pse*A*, his*H1*, his*F2*, pse*H*, pse*G, and *pse*F), a region known to be hypervariable in *Campylobacter* spp. [73]. This pathway synthesises and attaches pseudaminic acid to flagellar proteins and is important for motility, colonisation, and immune evasion [74, 75]. It is therefore plausible that large-scale changes to this pathway, attributed to the importation of this tract into Clade F_REC through natural transformation, have provided a competitive advantage, leading to the emergence and dominance of this lineage in both the poultry reservoir and human cases.

While the importation of a tract that resulted in major changes in flagellar glycosylation may have facilitated the emergence of Clade F_REC, the loss of the methionine biosynthesis pathway (*met*AB) through a separate recombination event may have contributed to its decline. Methionine biosynthesis involves multiple pathways, including a pathway involving *lux*S, which was present in all ST474 isolates, but experimental evidence showed that the absence of the *met*AB operon was associated with reduced, but did not prevent, colonisation in chickens [76]. Whether the loss of this pathway contributed to the decline of re-emergence Clade F remains highly speculative and would need to be supported by colonisation studies. More broadly, evidence from this study that the dynamics of ST474 and the poultry-associated epidemic and re-emergence clades E and F may be driven by genomic events, in addition to abiotic factors attributable to the interventions in the poultry industry, requires further investigation.

The presence of two integrated elements (IEs) in ST474 is consistent with other studies of *C. jejuni* highlighting the role of phage-mediated transduction in horizontal gene transfer. These prophage-derived islands have been characterised and classified [57]. Experimental studies have demonstrated their potential role in regulating natural transformation and stabilizing lineages through the action of endonucleases, such as the endogenous DNase identified in CJIE1 in our study [67, 77]. The presence of endonuclease and methyltransferase genes in CJIE5 suggest it may play a similar role in reducing the uptake of foreign DNA. IEs have also been reported to play a role in adherence and invasion of host cells [58, 59]. The emergence of AMR strain ST6964 in New Zealand was also characterised by the presence of multiple prophage homologues, with evidence of plasmid mediated introduction of an IE into the chromosome [78].

CJIE1 is a ∼40kb IE identified in a subset of isolates in all clades, including the ‘basal’ Clade A and other isolates belonging to CC48, the CC incorporating ST474. This is consistent with the vertical inheritance of CJIE1 from the common ancestor of all ST474 isolates, with subsequent loss in multiple clades potentially attributable to low levels of negative selection pressure. CJIE1 is commonly found in *Campylobacter jejuni*, including reference strain RM1221 [57]. If the IEs have contributed to lineage stability through the inhibition of natural transformation [67, 77] this may account for the relatively low proportion of the genome in recombinant regions in Clade A and E. Conversely, the loss of CJIE1 from multiple clades may have contributed to the increase in recombination observed in clades B-F, including Clade F_REC. Similarly, the loss of CJIE5 may have contributed to the relatively high levels of recombination in Clade F_REC.

CJIE5 was consistently located in an integration site between *cgp*A and *traF*. In contrast, CJIE1 was observed in multiple locations within the ST474 genomes, including directly upstream of CJIE5 in isolates in Clade E. Variants of CJIE1 have shown similar mobility in other *C. jejuni* STs, including the AMR New Zealand epidemic strain ST6964 [78]. CJIE5 was present in Clade A and poultry associated epidemic Clade E. Its sporadic presence in clades B-D suggests this IE may have been acquired from a common ancestor of ST474. Its absence in ST48 is consistent with the acquisition being more recent than the common ancestor of ST474 and ST48. In ST474 isolates, CJIE5 contained a 177bp gene at the right-end junction, containing multiple poly T and poly A tracts that was annotated as a tRNA-Leu due to the high sequence similarity with other tRNA-Leu genes. tRNA-Leu genes are associated with the site of insertion of chromosomally integrated elements in *C. jejuni* [56]. However, the CJIE5 identified in ST677 by Skarp et al [56], while adjacent to a tRNA-Leu at the left end junction, did not have the tRNA-Leu ‘pseudogene’ at the right end junction. tRNA-Leu genes are associated with both CJIE5 and CJIE2 in *Campylobacter* spp. The location of these IEs adjacent to these tRNAs is likely due to these regions being transcriptionally active.

Our understanding of the characteristics and role of plasmids in *Campylobacter* spp. is limited compared to other bacterial species. The three plasmids identified in this study were typical of conjugative plasmids and their location in the phylogeny of isolates in Clades B-D, and their presence in other isolates in the SACNZ study from diverse lineages, suggests they have been horizontally acquired on multiple occasions. None of the plasmids were observed in poultry-associated Clades E & F or basal Clade A. Plasmid type M4_R59_S3_V1 is globally associated with the presence of tetracycline resistance gene *tet*O, including the similar plasmid p18682, but in the SACNZ study only one isolate was confirmed as carrying this gene in a M4_R59_S3_V1 plasmid (belonging to ST45). Two other SACNZ isolates (ST50) carried *tet*O in a plasmid contig that carry one or more the markers belonging to M4_R59_S3_V1. This is consistent with the relatively low use of antimicrobials in agriculture in New Zealand [79], although the country did experience a major poultry-associated outbreak of campylobacteriosis associated with a tetracycline and fluoroquinolone resistant *C. jejuni* (ST6964) between 2014 and 2016 [78], in which the *tet*O gene was chromosomally located.

Although annotation tools such as Bakta assign *virB*/*trb* designations to several plasmid-borne genes identified in this study, it should be noted that these genes have not been confirmed as encoding a functional type IV secretion system (T4SS). The *virB* gene set on *Campylobacter* plasmids is incomplete relative to the T4SS reference *Agrobacterium tumefaciens* VirB/VirD4 system, and the operon is incomplete in *Campylobacter* plasmids [80]. The operon structure remains unresolved, and no studies have confirmed a functional conjugation system similar to T4SS in *Campylobacter* plasmids [81].

The importance of the bacterial accessory genome in facilitating adaptation to new niches has been observed in many different bacterial species. A study of the emergence of *Enterococcus faecium* identified that not only were plasmids critical for the acquisition of AMR, they were also more informative for source attribution than chromosomes [82]. Plasmids were also the vehicle for adaptation of *Salmonella* in Brazilian poultry in response to vaccine introduction and increasing antibiotic selection pressure [83]. Other mobile genetic elements also play a role in enabling pathogens to expand their niche; in monophasic *Salmonella* Typhimurium ST34, acquisition of a novel prophage encoding AMR genes and a genomic island encoding metal tolerance genes likely helped ST34 become one of the most common *Salmonella* subtypes globally, found across multiple host species and countries [84].

Source attribution using genomic information has traditionally used a few housekeeping genes (e.g. 7 gene MLST), where the genetic diversity is relatively low and thus sequence types among isolates found in the target population are normally also found among isolates from putative sources. In this scenario, count based models [85, 86] are useful for performing source attribution. When higher genomic precision is utilised, however, all isolates typically differ from each other, so that counts of each sequence observed are 1, with the number of each ‘target’ type observed on the sources typically being zero [85]. Thus, alternate models that utilise the genetic similarity between isolate types make more sense. The asymmetric island model utilises the allelic profile information (i.e. the counts used are the counts of each allele at each loci) with an evolution model that allows one allelic profile to transform to another via mutation (a single locus change to a new allele), recombination (combining previously observed alleles), or a combination thereof [87]. Thus, the asymmetric island model allows attribution even in the case of wgMLST data.

The role of ST474 in the epidemic of *Campylobacter* in New Zealand has previously been described for the time period 2005-2008, where a significant decline in cases was described following food safety interventions in the poultry industry [6]. These earlier studies were based on low resolution 7-gene MLST. With longitudinal high resolution genomic data, we were able to explore the dynamics of this strain in much greater detail over a longer time period.

We can speculate from the genomics and modelling that the emergence of epidemic Clade E was preceded by an ancestral host jumping event, followed by clonal expansion in a rapidly growing poultry industry [88, 89]. Meanwhile, other more stable lineages persisted in environmental reservoirs, including ruminants, as well as poultry. We have identified potential extrinsic and intrinsic drivers for the emergence of poultry-associated clades E and F and the decline of Clade F – including the role of transduction and natural transformation (and their interaction). However, it is not clear how such factors could have affected the entire reservoir of Clade E across multiple flocks of birds on multiple farms in different locations. The interventions were largely focussed on the processing plants [6], and it is unlikely these would have impacted the transmission dynamics at flock level. This raises the currently unanswered question: what extrinsic factors could have driven the apparent extinction of all lineages of both poultry-associated clades, particularly the epidemic Clade E? Understanding the reasons for this phenomenon could help with the design and implementation of future pre-processing interventions.

## Data Availability

All data produced in the present study are available upon reasonable request to the authors. All ST474 raw reads and assemblies generated for this analysis are available from NCBI (Bioproject ID: PRJNA1538059).

## Acknowledgements

DAW and NPF were supported by the New Zealand Food Safety Science and Research Centre (NZFRSSC). AEM is supported by Biotechnology and Biological Sciences Research Council (BBSRC) Institute Strategic Programme Microbes and Food Safety BB/X011011/1 and its constituent project BBS/E/F/000PR13634 (Theme 1, Microbial threats from foods in established and evolving food systems). Additional support was provided by BBSRC grant BB/V018221/1. We also acknowledge the initial funding and support for the campylobacteriosis sentinel surveillance site from the Ministry for Primary Industries, the poultry industry, and other members of the *^m^*EpiLab team who assisted with the collection and processing of samples.

## Ethical approval

Approval from the Multi-Region Ethics Committee, New Zealand, was provided for work related to the *Campylobacter* sentinel surveillance site (application no. MEC/10/16/EXP). Isolates were collected and analysed through routine public health investigation activities. Ethics approval for the SACNZ study was obtained from the Northern B Health and Disabilities Ethics Committee (HDEC).

## Supplementary Figures and Tables

**Supplementary Figure S1.**
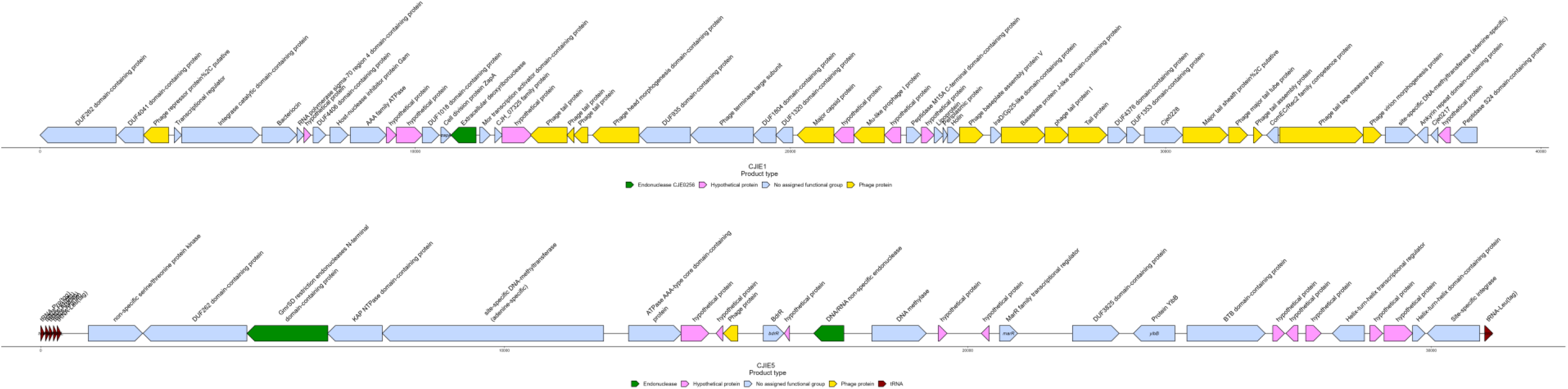

**Supplementary Figure S2a.**
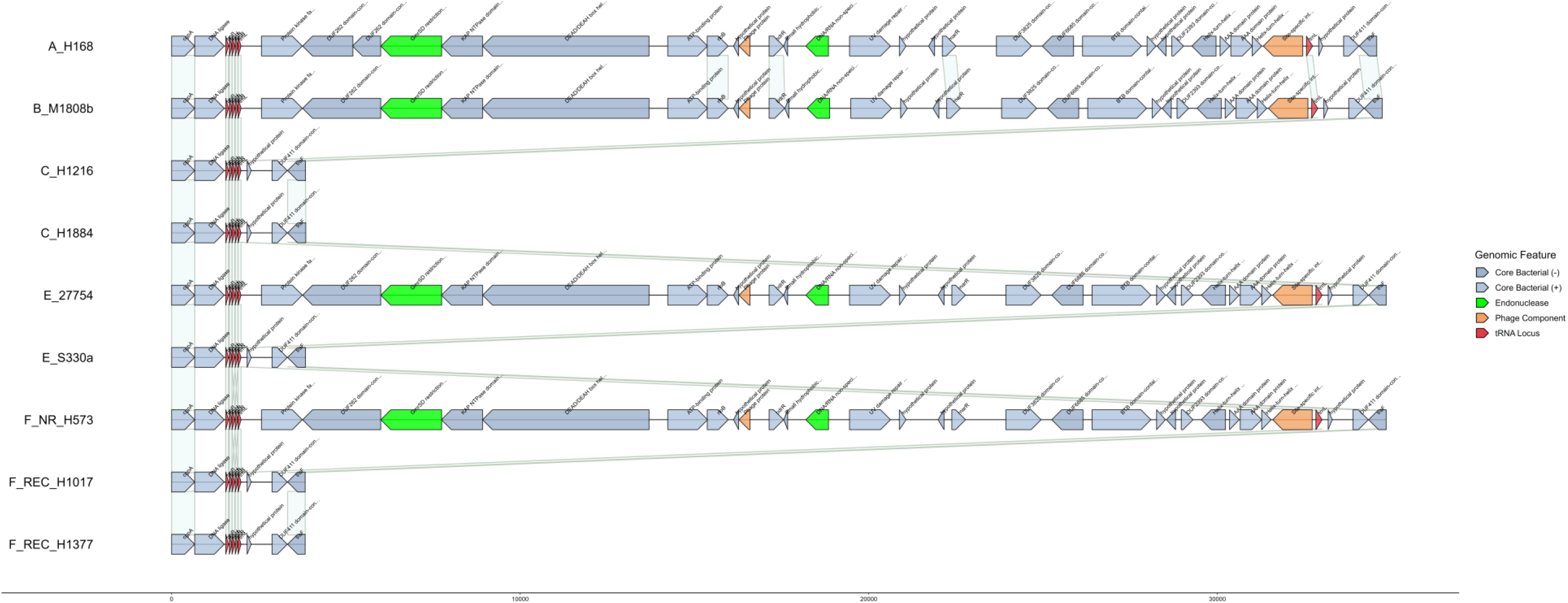
Insertion site *cgp*A – *traF* in 9 genomes belonging to multiple clades. Isolates A_H168, B_M1808b, E_27754 and F_NR_H573 all have CJIE5 inserted at the same location, the remaining isolates have the ‘empty’ insertion site with no integrated element.

**Supplementary Figure S2b.**
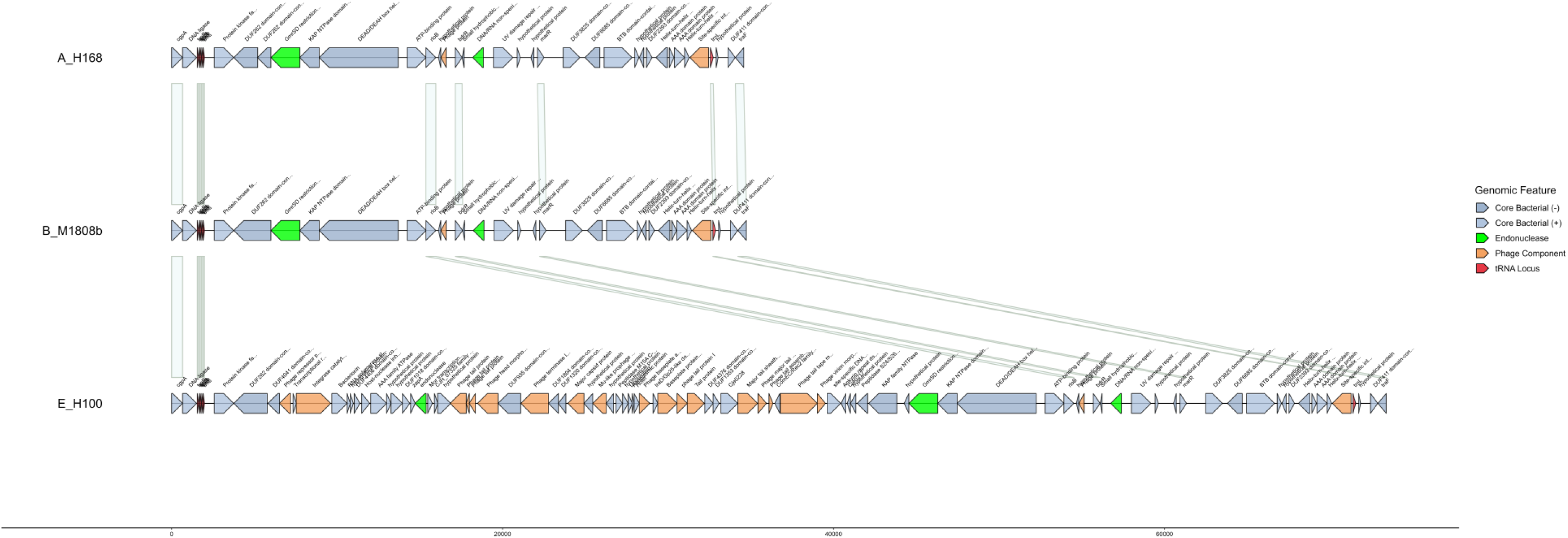
Integration site *cgp*A – *traF* comparing two genomes with CJIE5 (A_H168 and B_M1808b) and one with CJIE5 and CJIE1 (E_H100). Isolate E_H100 shows the presence of CJIE1 inserted into the *cgp*A – *traF* integration site, between the larger endonuclease and two to three CDSs downstream of the smaller tRNA-Leu in CJIE5.

**Supplementary Figure S2c.**
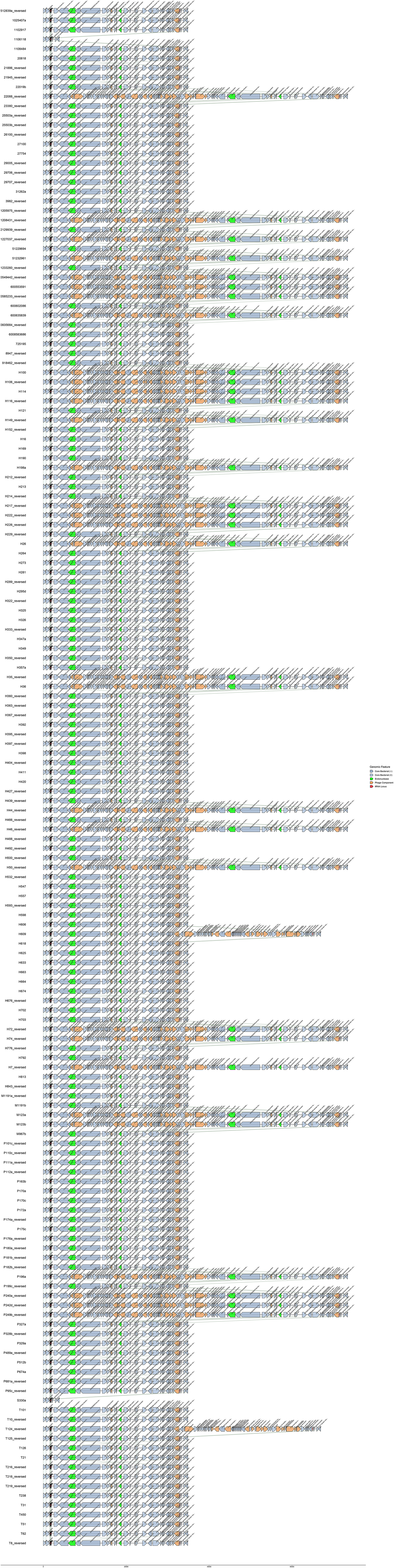
Integration site *cgp*A – *traF* for all ST474 Clade E isolates with the full insertion site on a single contig (N=162).

**Supplementary Figure S3.**
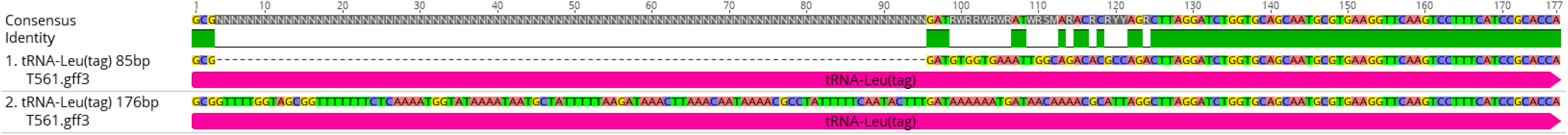
Alignment of the 85bp tRNA-Leu and the 177bp gene annotated as a tRNA-Leu.

**Supplementary Figure S4.**
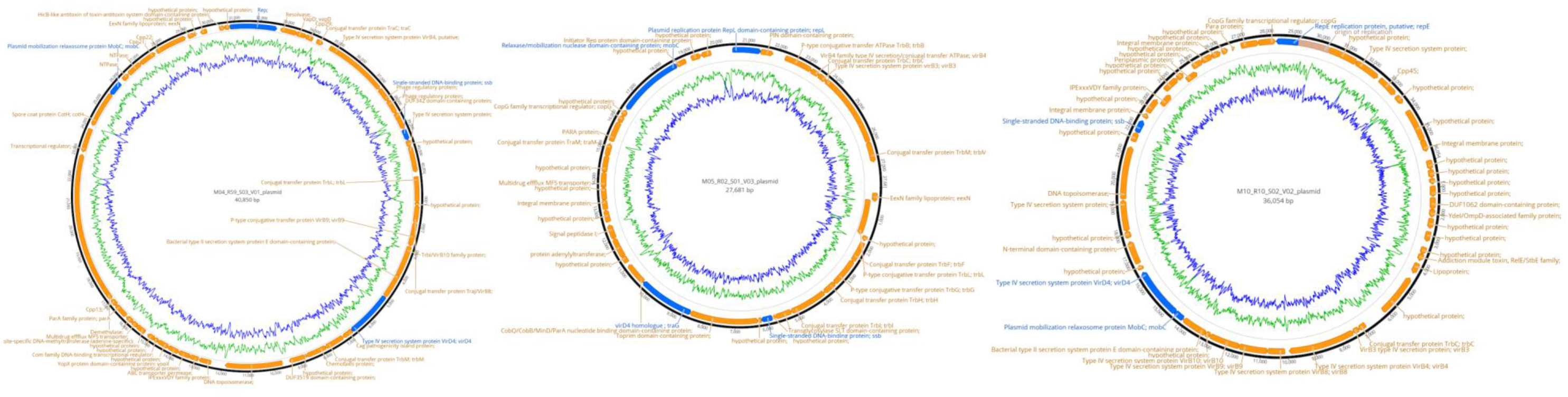
Comparison of the three plasmids identified in the ST474 in this study, showing genes and putative products.

**Supplementary Figure S5:**
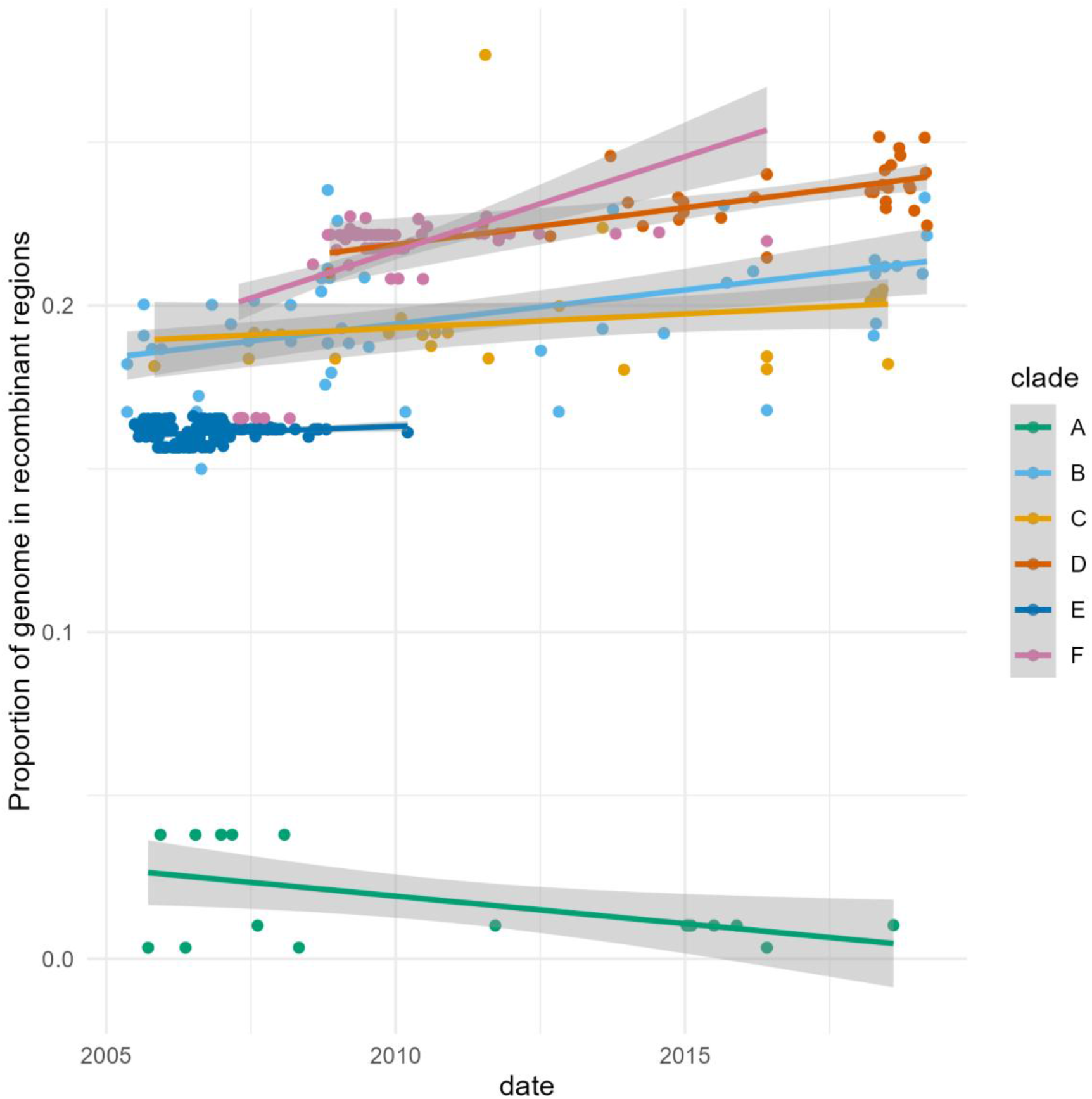
Linear regression of the proportion of genomic content included in recombinant regions over time (date of isolation). Data coloured by clade.

**Supplementary Figure S6.**
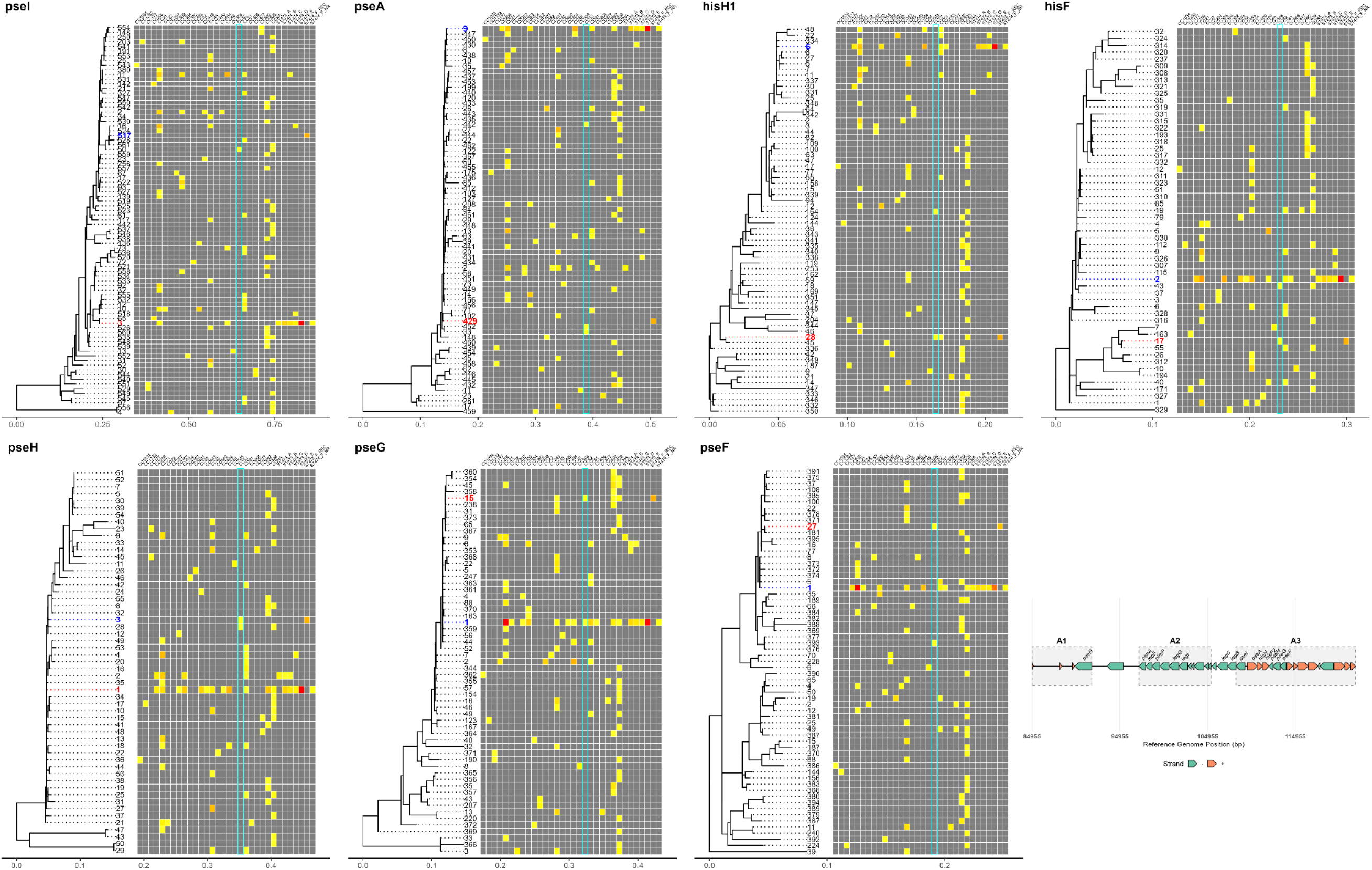
Maximum likelihood phylogenies of seven genes in the pseudaminic and legionaminic acid biosynthesis pathways, involved in flagellar glycosylation in *Campylobacter* spp. The blue tip label corresponds to the allele most prevalent in ST474 Clades A-E and Clade F_NR. The red tip label corresponds to the allele identified in Clade F_REC. The heatmap shows the alleles present in each ST474 clade and Clonal Complexes identified in the SACNZ study, with grey representing not present, red being the most frequent and yellow being the least frequent. The cyan box highlights the alleles identified in CC52, which shares alleles with ST474 Clade F_REC. The coordinates of genes in recombinant Regions A1-A3 are shown (reference genome H1216). The alleles in recombinant Clade F_REC were phylogenetically distinct from those in isolates in other ST474 isolates. Their similarity with alleles from different CCs indicates an importation of genes from an isolate or isolates belonging to a divergent lineage.

**Supplementary Figure S7.**
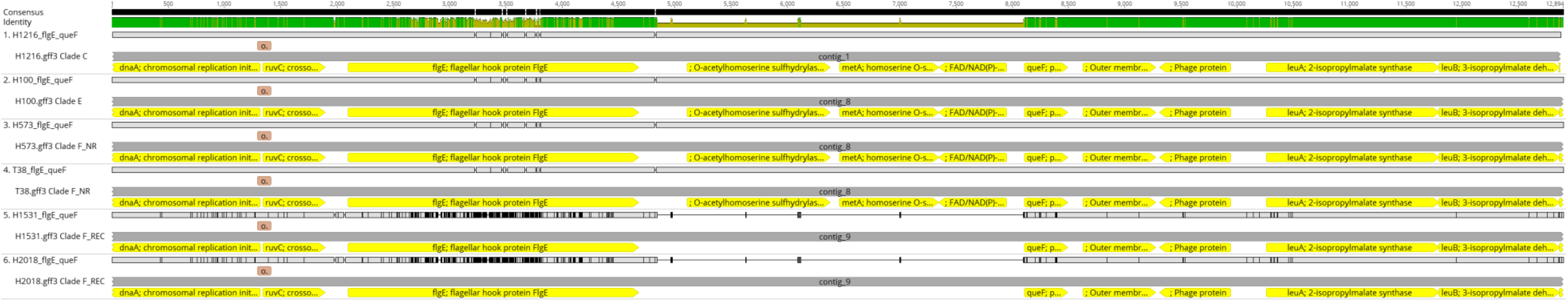
Alignment of the *met*AB locus and flanking genes in recombinant region F. H1216 is the reference and belongs to Clade C, H100 belongs to Clade E, H573 and T38 belong to Clade F_NR. The bottom two isolates belong to clade F_REC (H1531 and H2018) and show the deletion of the *met*AB operon. *met*B is labelled O-acetylhomoserine sulfhydrylase. Evidence that the region including flanking genes flgE and queF and intergenic regions was subject to a recombination event is further supported by the large number of SNPs in these loci.

**Supplementary Figure S8.**
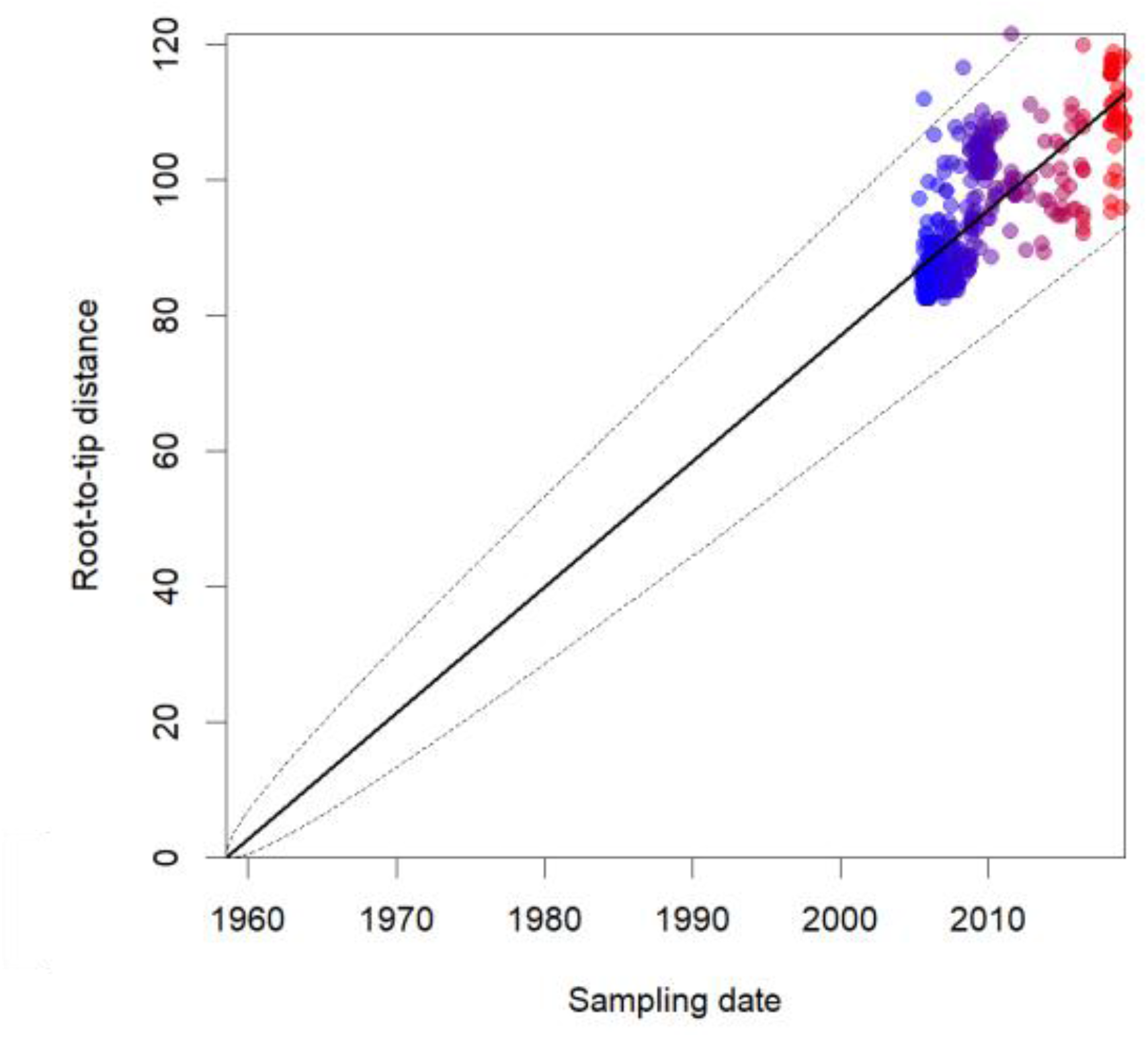
Root to tip regression of 443 ST474 isolates. The red dots are more recent samples and the blue dots are older samples.

**Supplementary Figure S9.**
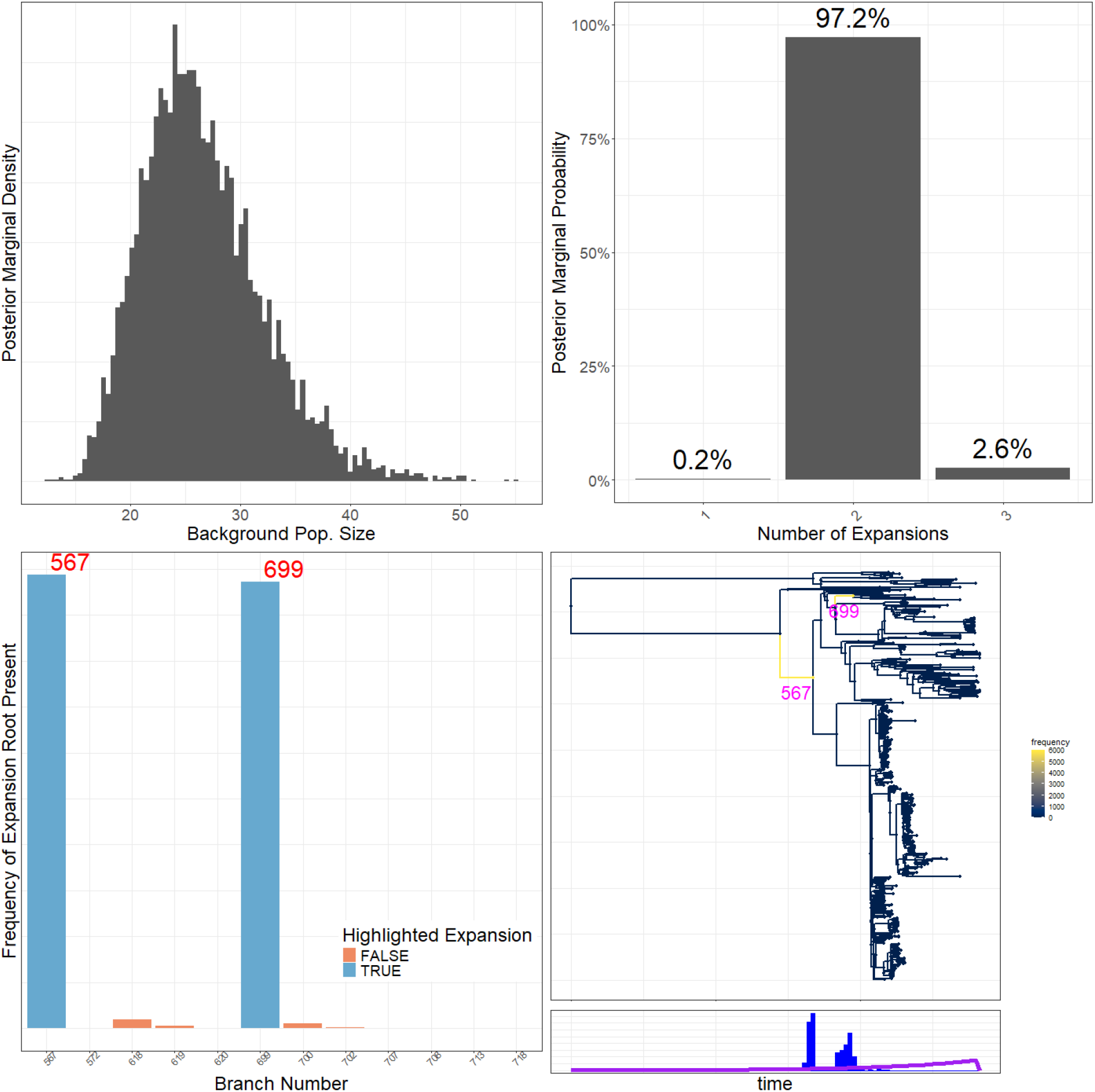
CaveDive summary plot showing support for the number of clonal expansions (top right and bottom left). The branches are identified in the tree (bottom right).

**Supplementary Figure S10.**
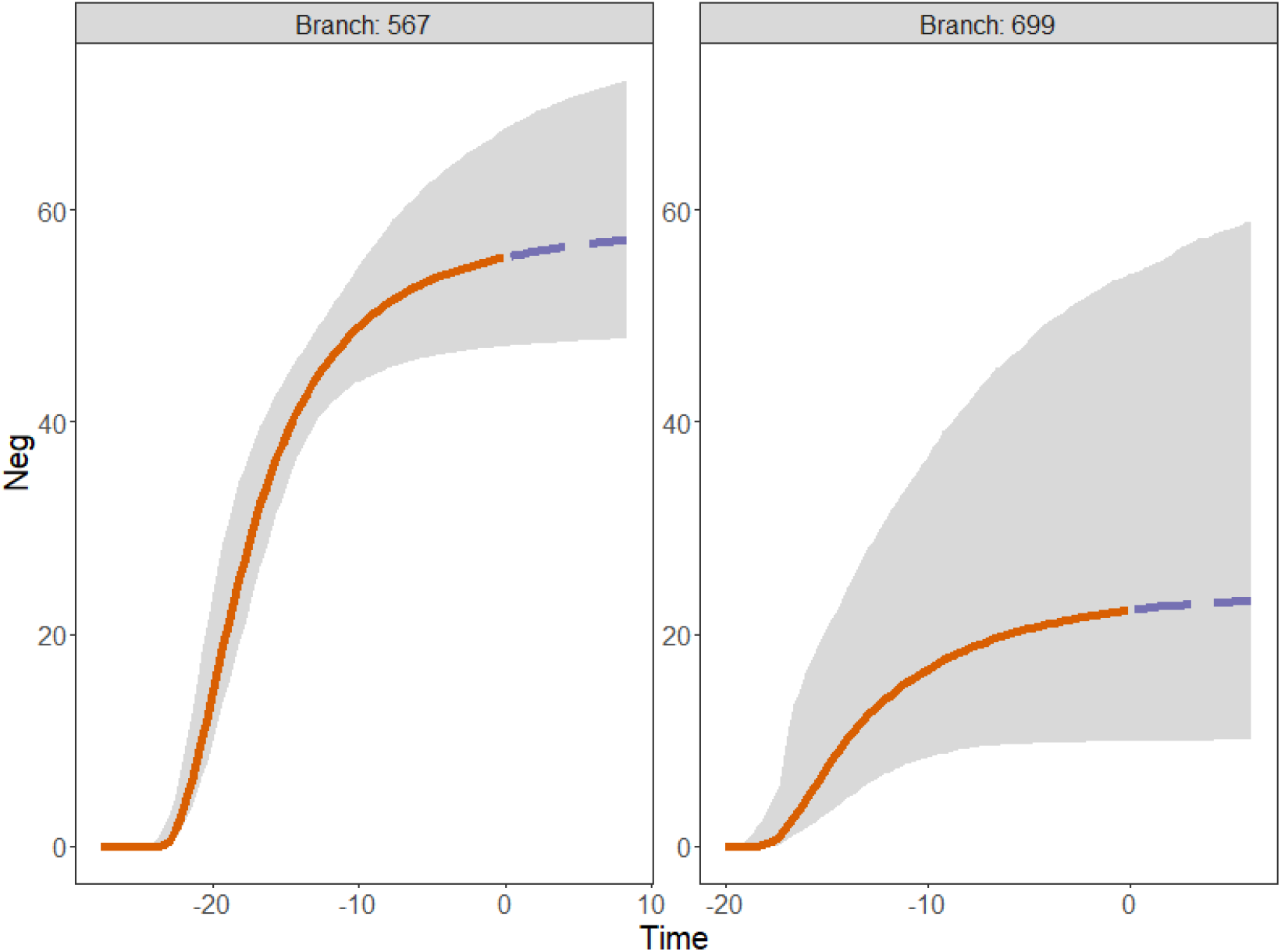
CaveDive plot showing estimated effective population sizes for the two branches with evidence of clonal expansion shown in red in Figure 3a. The x axis is the time in years from the most recent sample.

**Supplementary Table S1.**
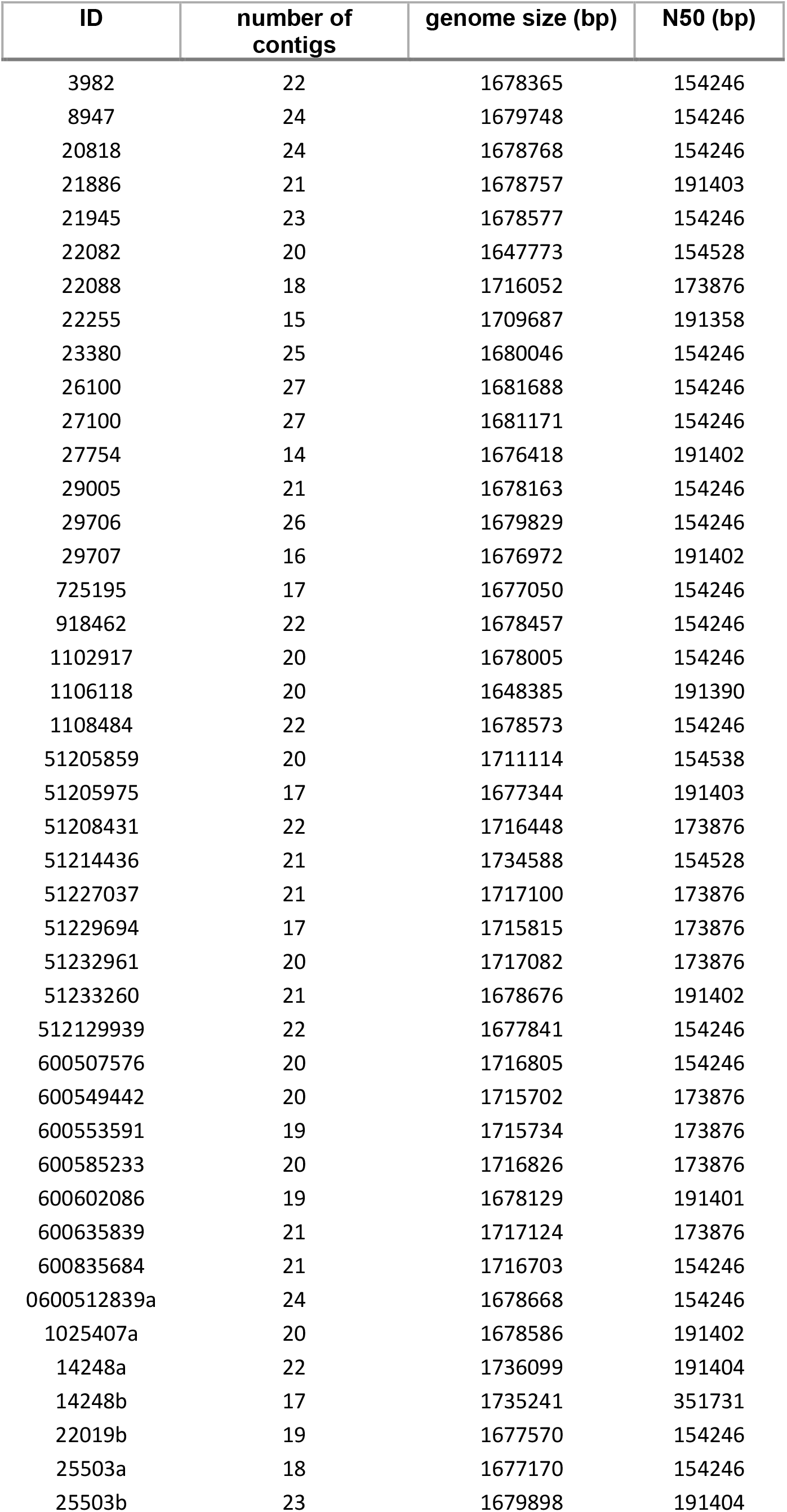
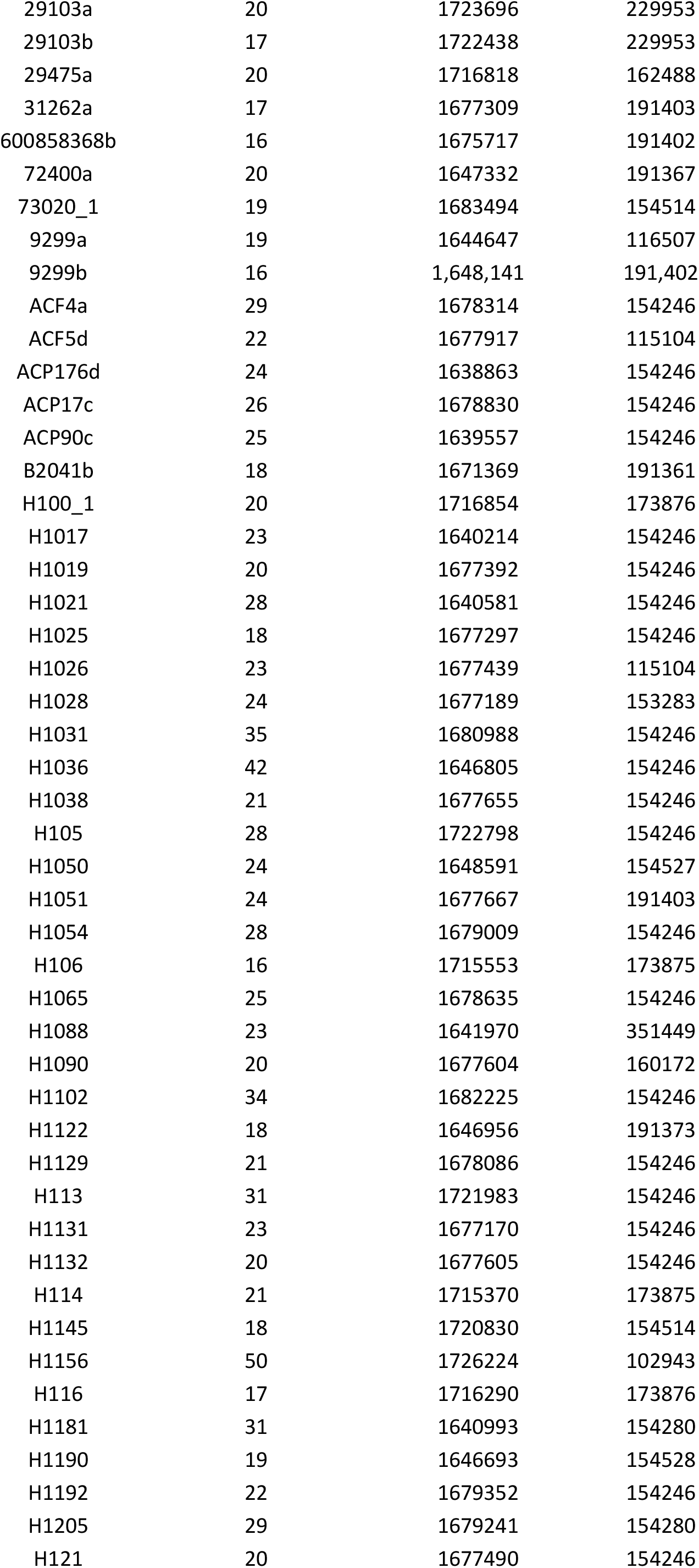
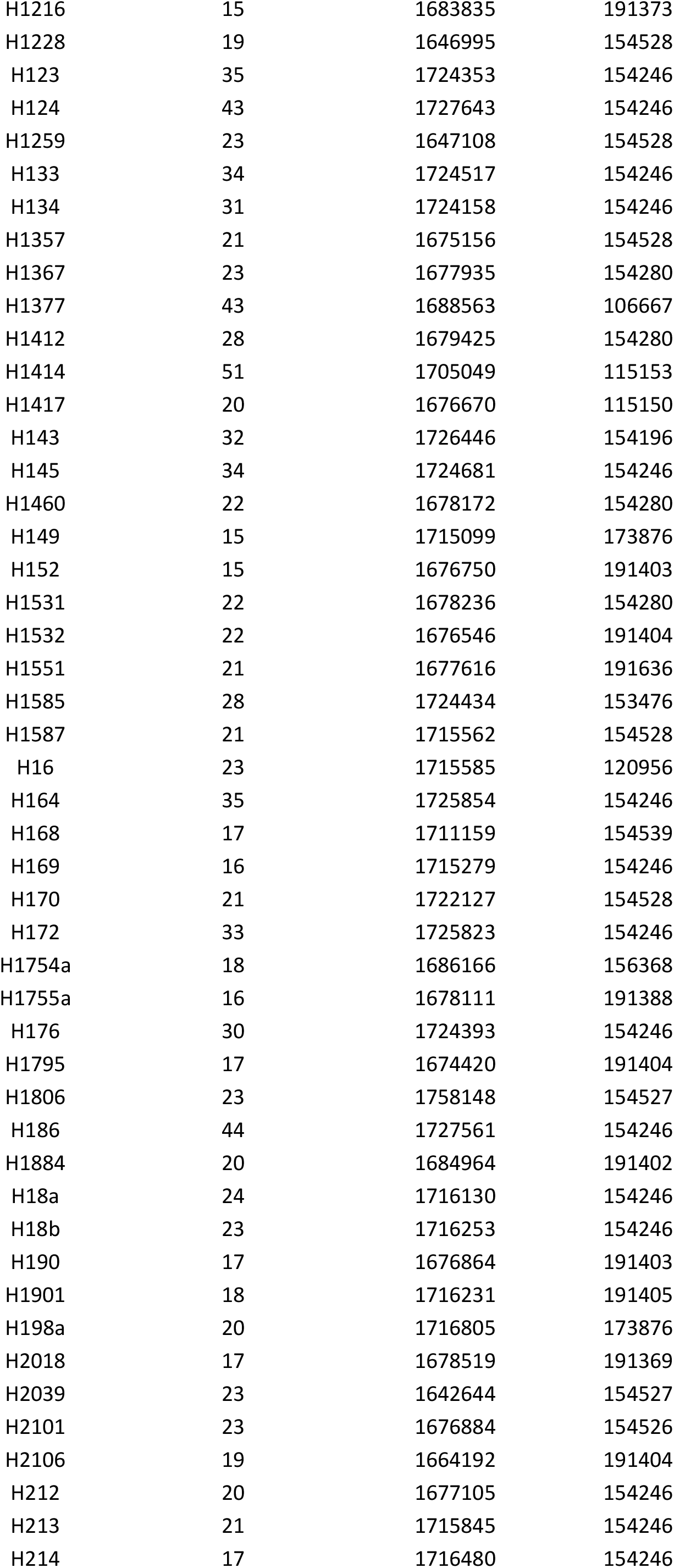
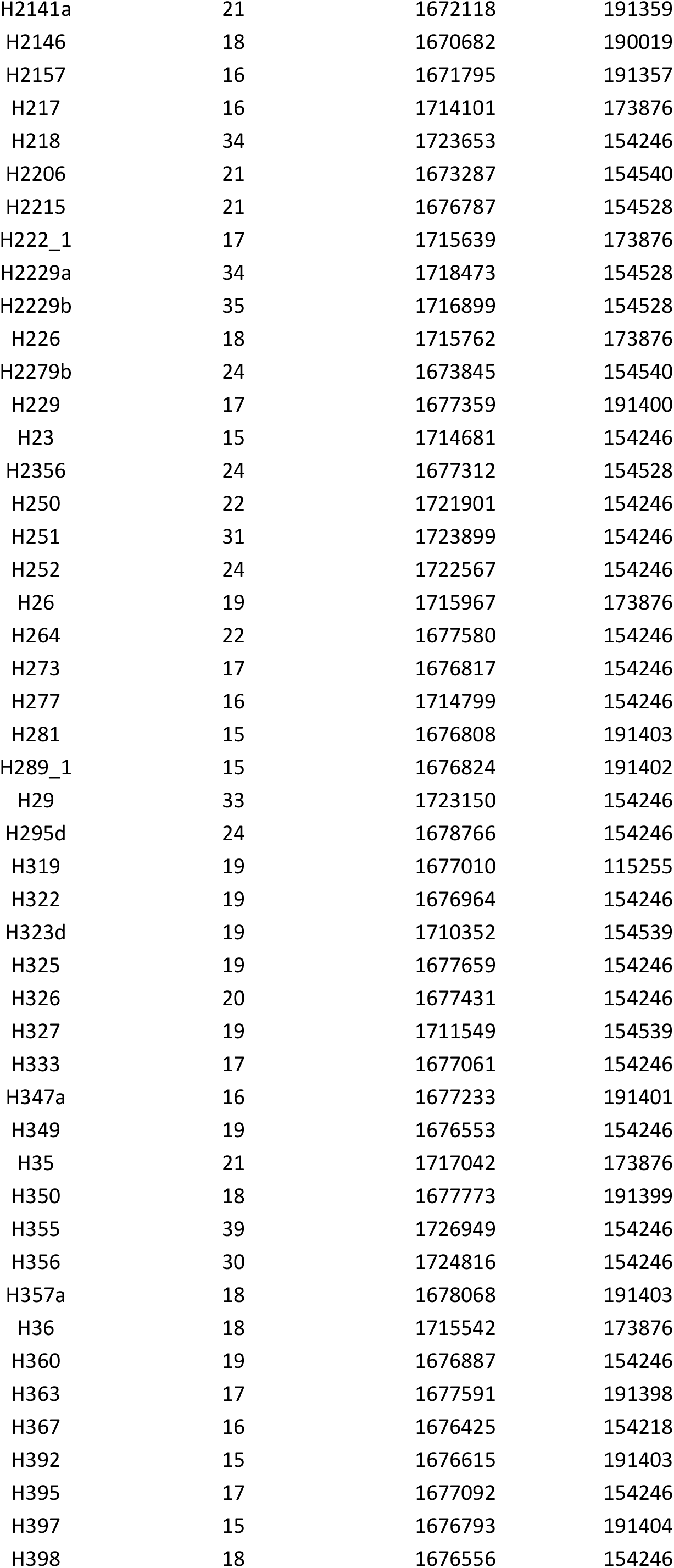
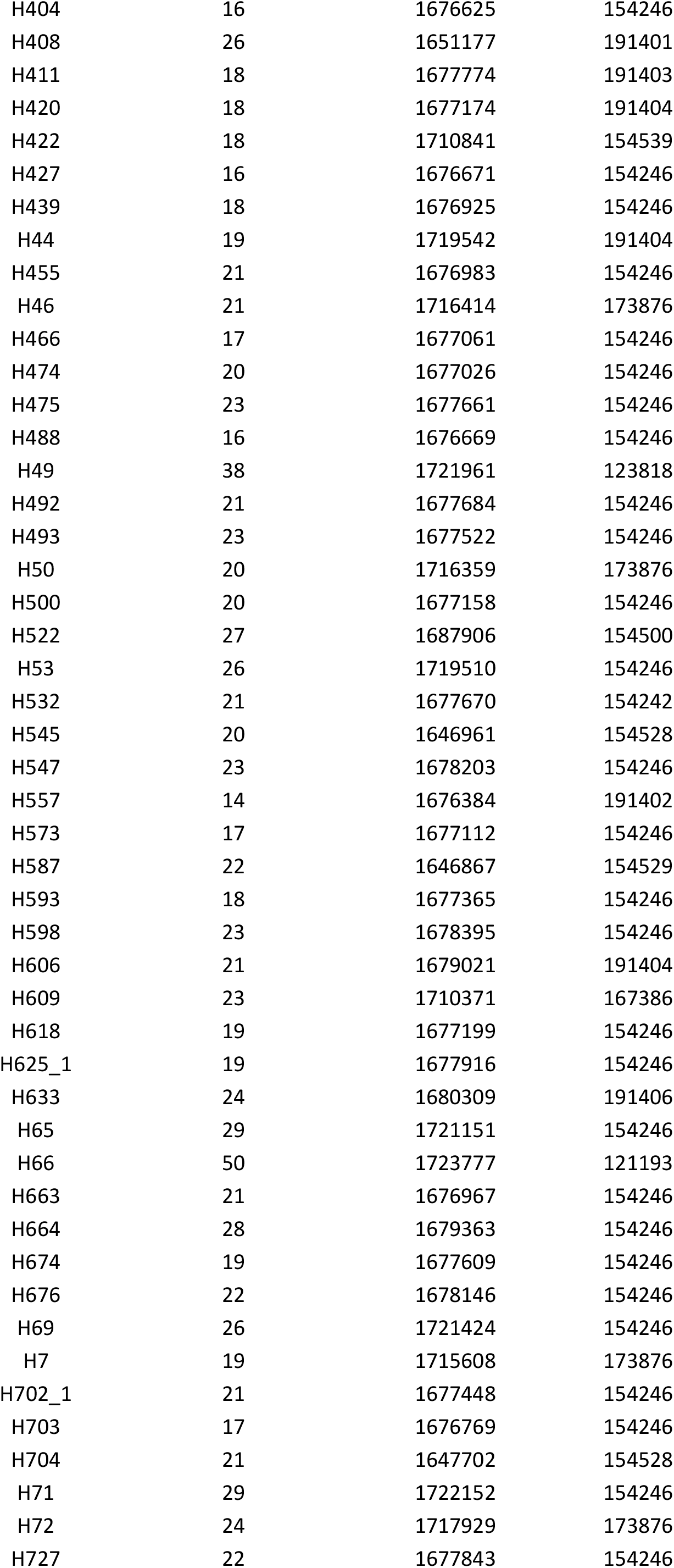
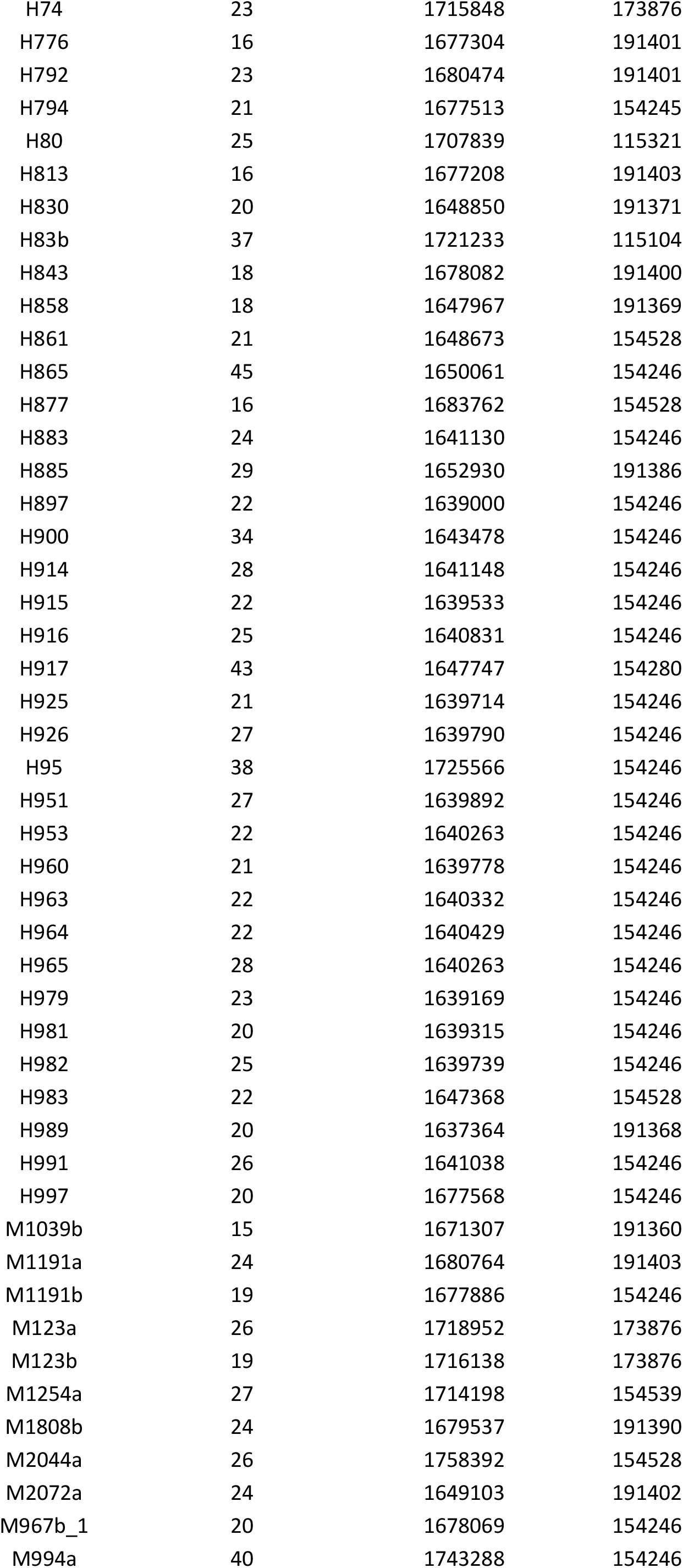
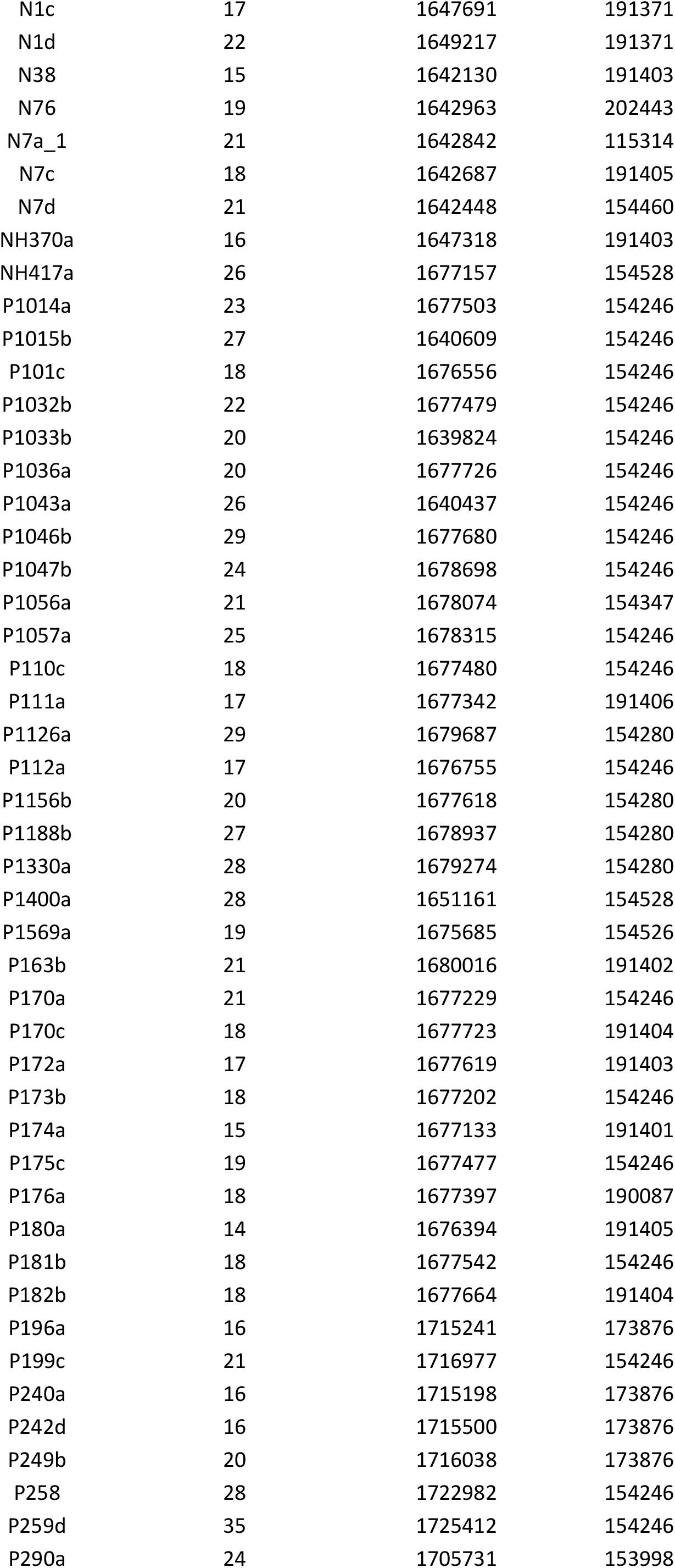
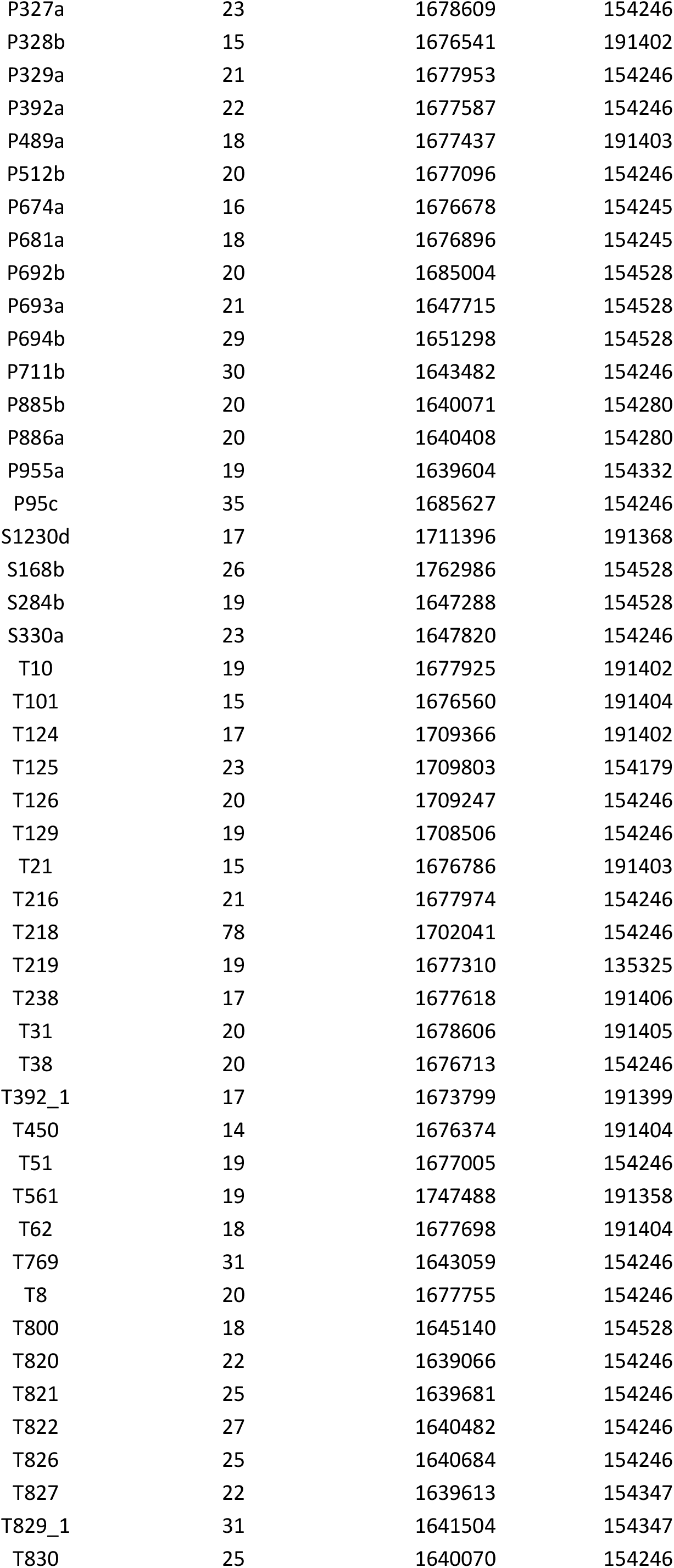
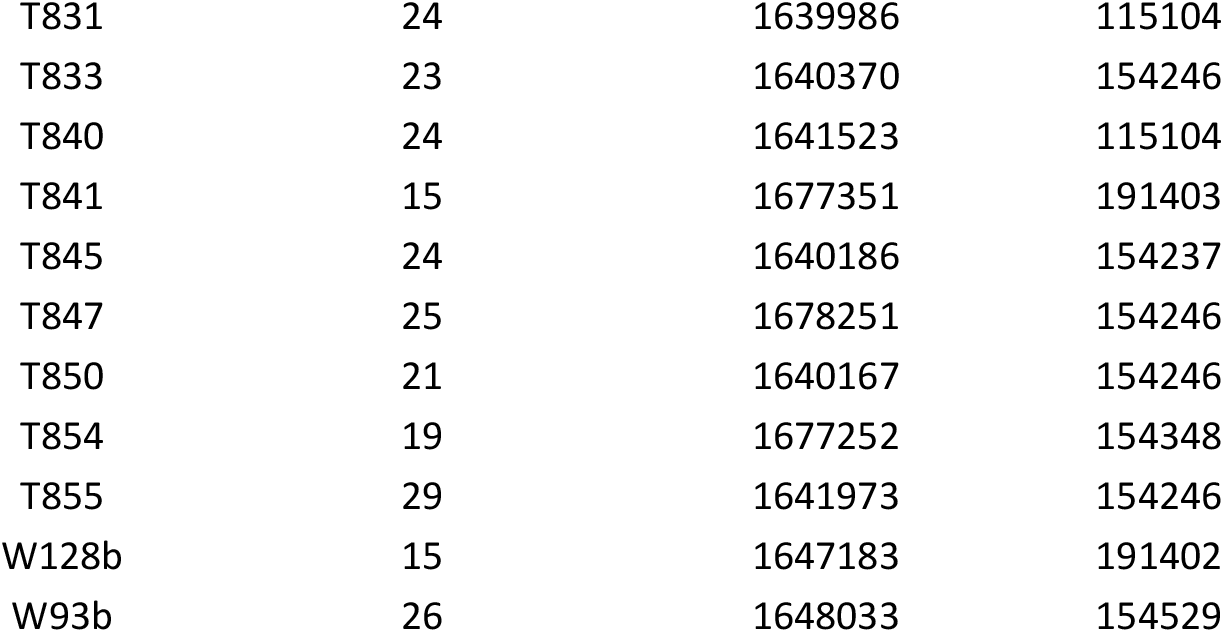
Assembly statistics for all 390 *Campylobacter jejuni* ST474 isolates generated as part of this study.

**Supplementary table 2.**
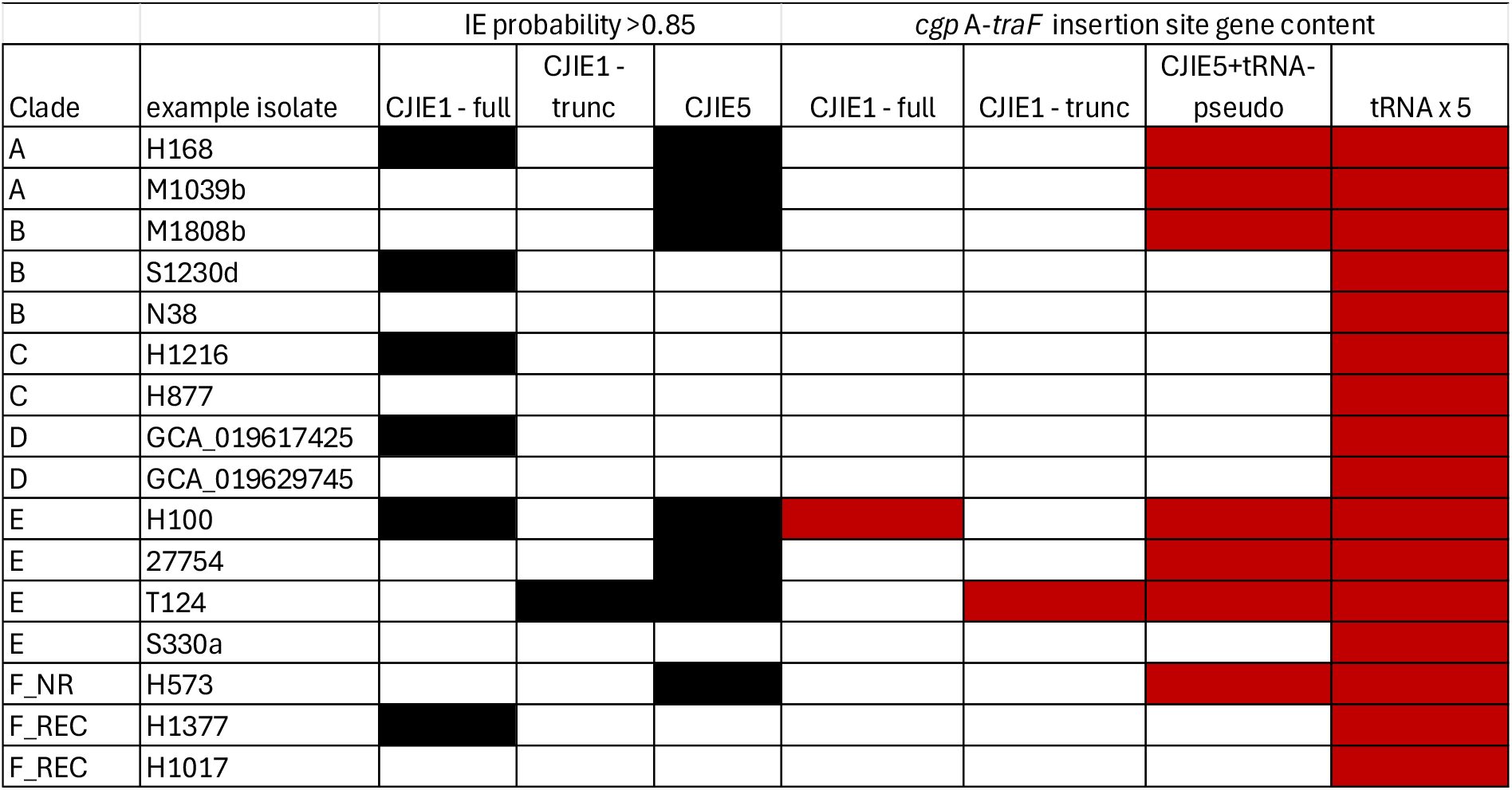
Contents of hypervariable integration site *cgpA-traF*. Black boxes indicate presence of the integrated element in the isolates, and red boxes indicate gene presence in the integration site.

**Supplementary Table S3.**
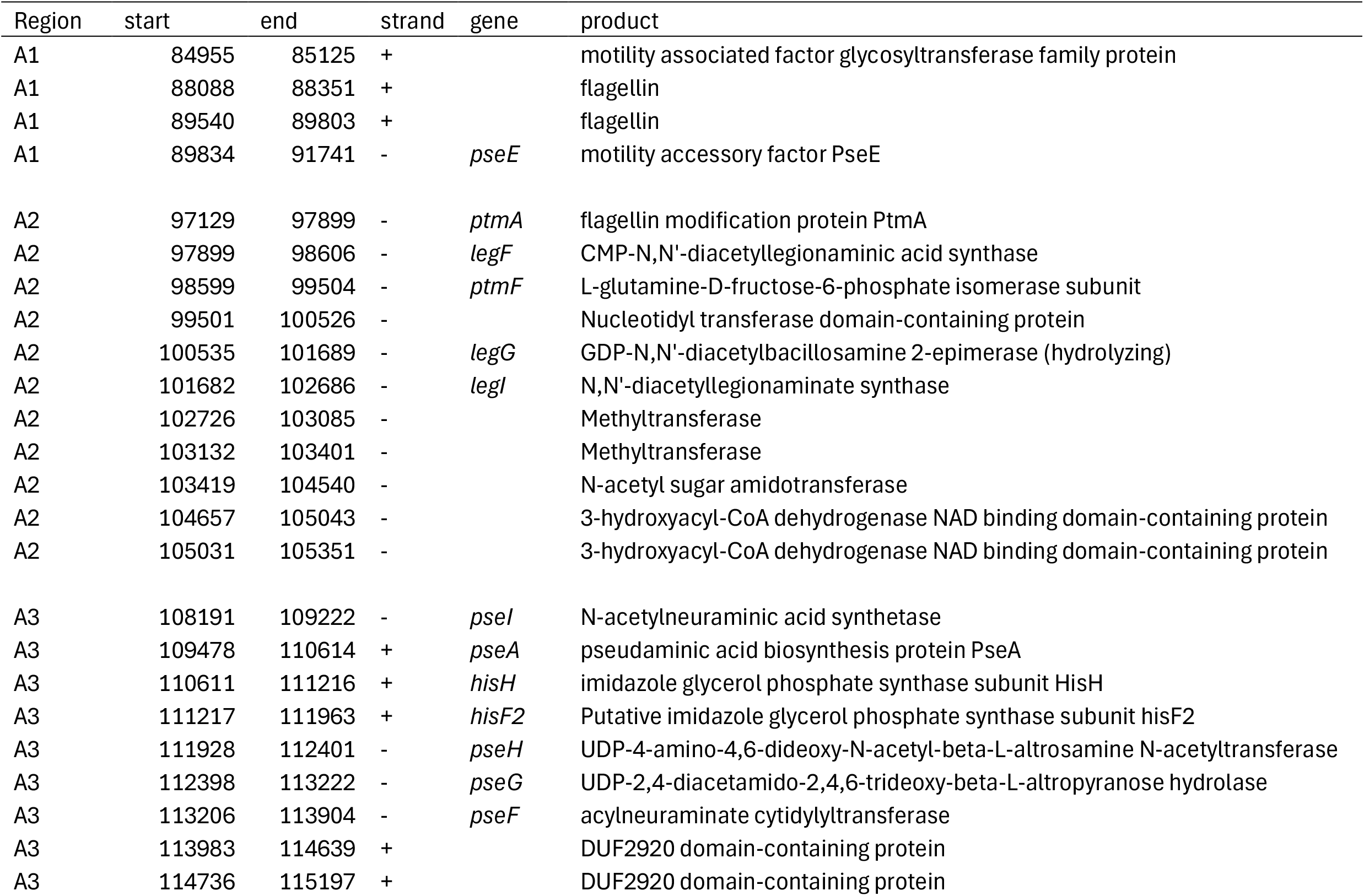
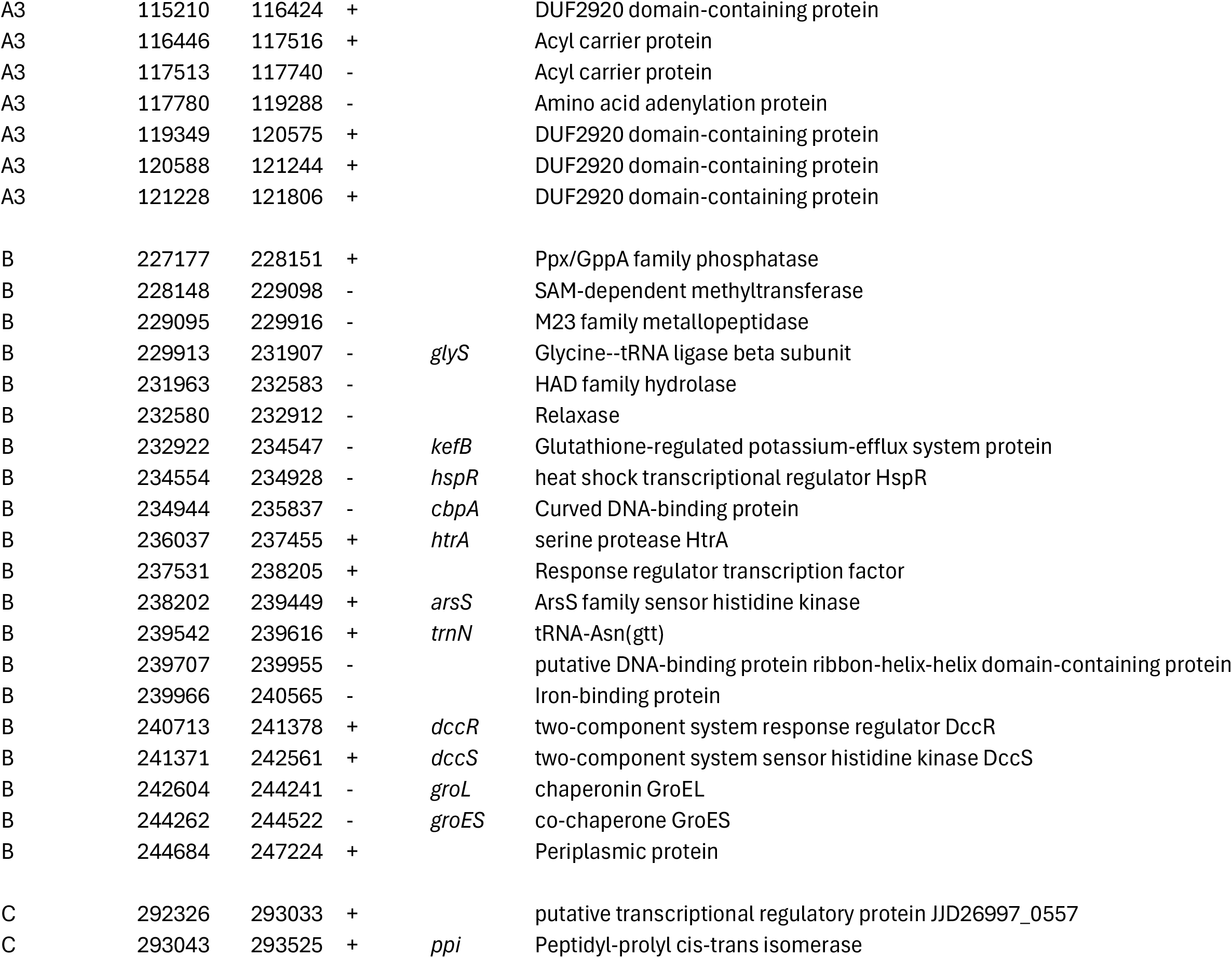
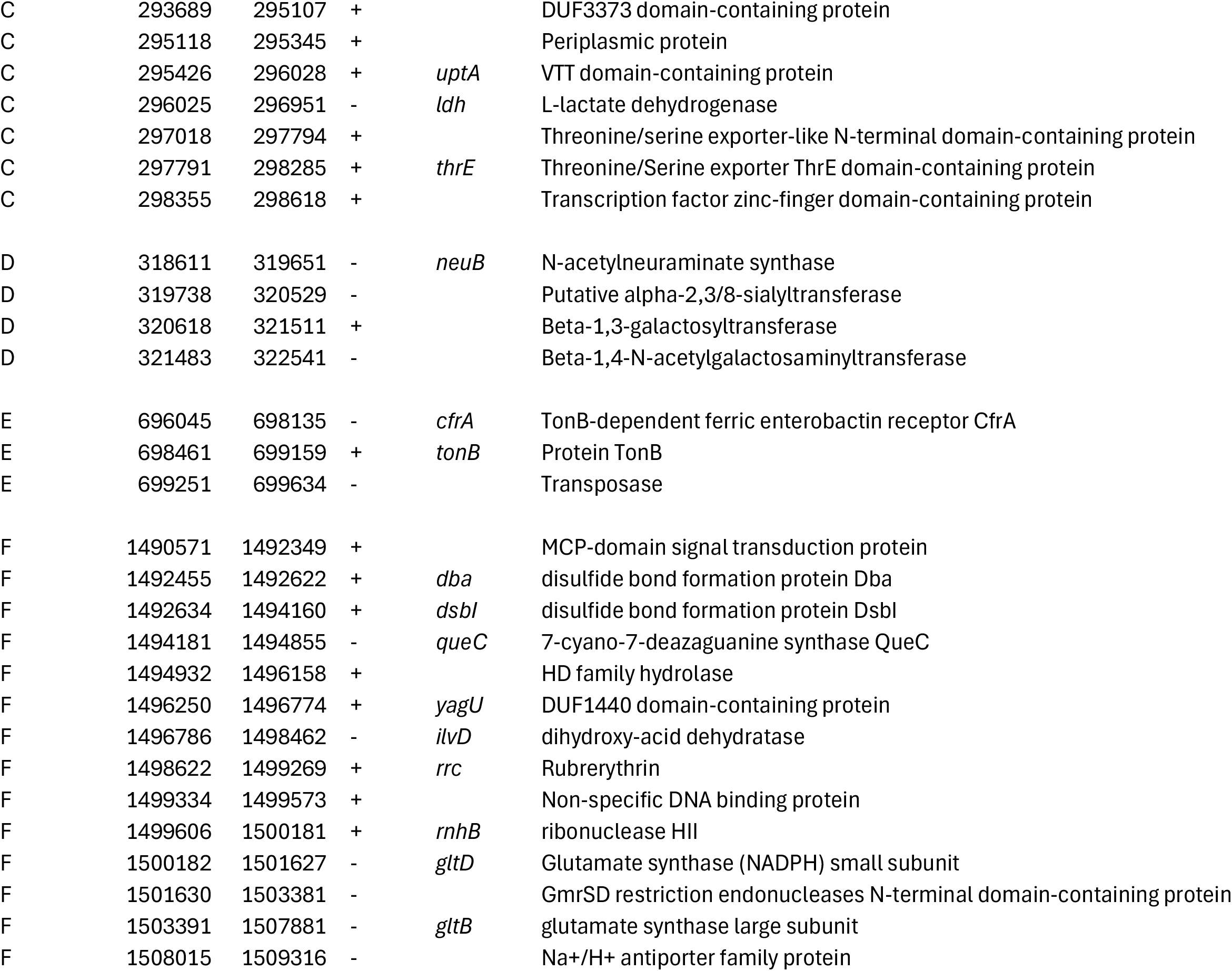
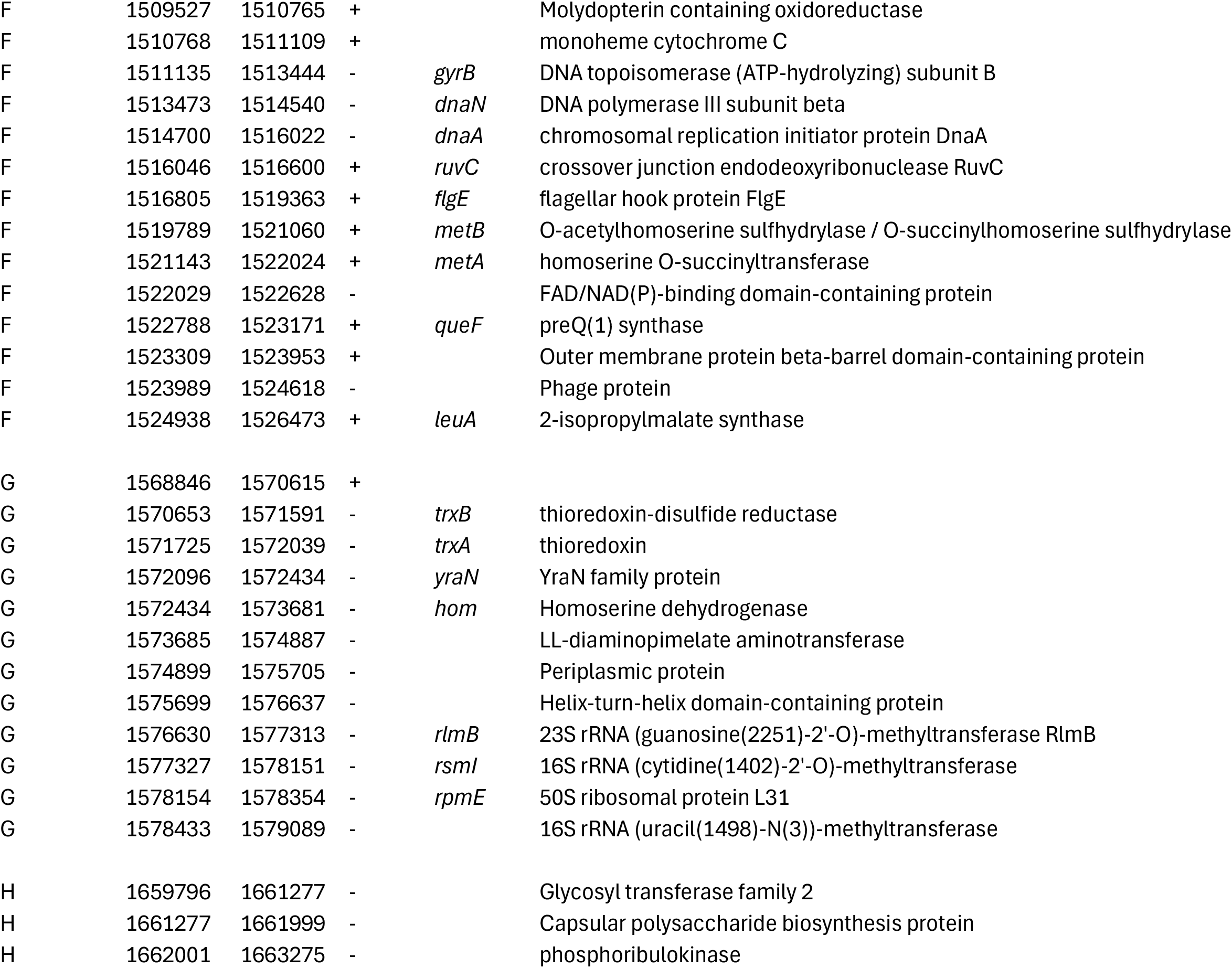
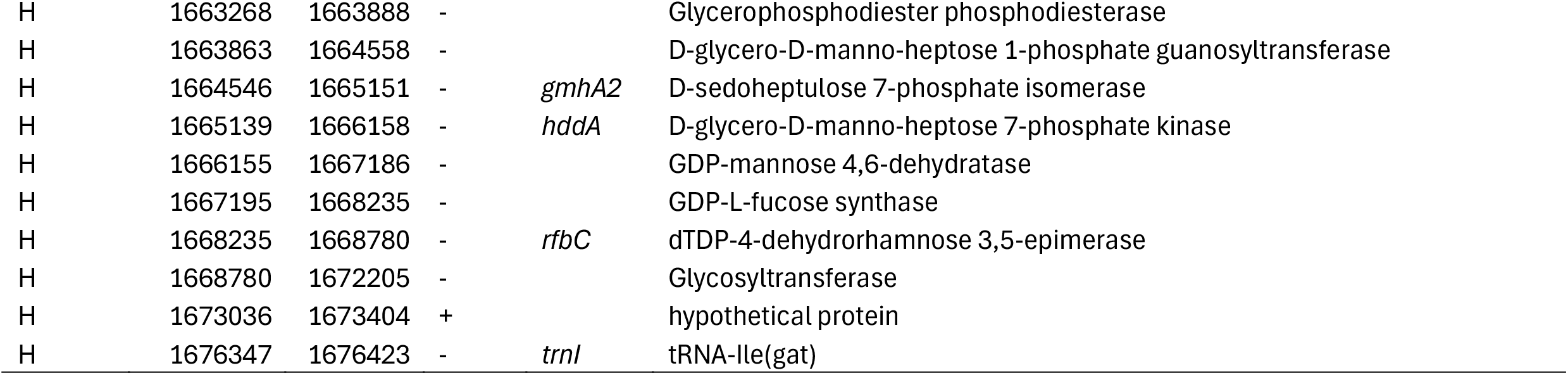
Gene content in regions identified as recombinant in Clade F.

## Footnotes

1 https://github.com/nickjcroucher/gubbins/blob/master/docs/gubbins_plotting.md

